# The polarity and specificity of SARS-CoV2-specific T lymphocyte responses determine disease susceptibility

**DOI:** 10.1101/2021.06.18.21258477

**Authors:** Jean-Eudes Fahrner, Agathe Carrier, Lyon COVID study group, Eric De Sousa, Damien Drubay, Agathe Dubuisson, Arthur Geraud, Anne-Gaëlle Goubet, Gladys Ferrere, Yacine Haddad, Imran Lahmar, Marine Mazzenga, Cléa Melenotte, Marion Picard, Cassandra Thelemaque, Luigi Cerbone, Joana R. Lérias, Ariane Laparra, Alice Bernard, Benoît Kloeckner, Marianne Gazzano, François-Xavier Danlos, Safae Terrisse, Carolina Alves Costa Silva, Eugenie Pizzato, Caroline Flament, Pierre Ly, Eric Tartour, Lydia Meziani, Abdelhakim Ahmed-Belkacem, Makoto Miyara, Guy Gorochov, Fabrice Barlesi, Caroline Pradon, Emmanuelle Gallois, Fanny Pommeret, Emeline Colomba, Pernelle Lavaud, Eric Deutsch, Bertrand Gachot, Jean-Philippe Spano, Mansouria Merad, Florian Scotte, Aurélien Marabelle, Frank Griscelli, Jean-Yves Blay, Jean-Charles Soria, Fabrice Andre, Mathieu Chevalier, Sophie Caillat-Zucman, Florence Fenollar, Bernard La Scola, Guido Kroemer, Markus Maeurer, Lisa Derosa, Laurence Zitvogel

**Affiliations:** Université Paris-Saclay, Faculté de Médecine, Le Kremlin-Bicêtre, France; Gustave Roussy, Villejuif, France; Institut National de la Santé et de la Recherche Médicale, UMR1015, Gustave Roussy, Villejuif, France; Transgene S.A., Illkirch-Graffenstaden, France; Open Innovation & Partnerships (OIP), bioMérieux S.A., Marcy l’Etoile, France. R&D – Immunoassay, bioMérieux S.A., Marcy l’Etoile, France; Joint Research Unit Hospices Civils de Lyon-bioMérieux, Civils Hospices of Lyon, Lyon Sud Hospital, Pierre-Bénite, France; International Center of Research in Infectiology, Lyon University, INSERM U1111, CNRS UMR 5308, ENS, UCBL, Lyon, France; Hospices Civils de Lyon, Lyon Sud Hospital, Pierre-Bénite, France; Département d’Oncologie Médicale, Gustave Roussy, Villejuif, France; Département d’Innovation Thérapeutique et d’Essais Précoces (DITEP), Gustave Roussy, Villejuif, France; Aix-Marseille Université, Institut de Recherche pour le Développement, Assistance Publique – Hôpitaux de Marseille, Microbes Evolution Phylogeny and Infections, Marseille, France; Département de Biostatistique et d’Epidémiologie, Gustave Roussy, Université Paris-Saclay, Villejuif, France; ImmunoTherapy/ImmunoSurgery, Champalimaud Centre for the Unknown, Lisboa, Portugal; Department of Immunology, Hôpital Européen Georges Pompidou, AP-HP, Paris, France; PARCC, INSERM U970; Univ Paris Est Creteil, INSERM U955, IMRB, Creteil, France; Sorbonne Université/Institut National de la Santé et de la Recherche Médicale, U1135, Centre d’Immunologie et des Maladies Infectieuses, Hôpital Pitié-Salpêtrière, Assistance Publique – Hôpitaux de Paris, Paris, France; Aix Marseille University, CNRS, INSERM, CRCM, Marseille, France; Centre de ressources biologiques, ET-EXTRA, Gustave Roussy, Villejuif, France; Département de Biologie Médicale et Pathologie Médicales, service de biochimie, Gustave Roussy, Villejuif, France; Département de Biologie Médicale et Pathologie Médicales, service de microbiologie, Gustave Roussy, Villejuif, France; Département Interdisciplinaire d’Organisation des Parcours Patients, Gustave Roussy, Villejuif, France; Institut National de la Santé et de la Recherche Médicale, U1030, Gustave Roussy, Villejuif, France; Département de Radiothérapie, Gustave Roussy, Villejuif, France; Service de Pathologie Infectieuse, Gustave Roussy, Villejuif, France; Department of Medical Oncology, Pitié-Salpétrière Hospital, APHP, Sorbonne Université, Paris, France; Service de médecine aigue d’urgence en cancérologie, Gustave Roussy, Villejuif, France; Center of clinical investigations BIOTHERIS, INSERM CIC1428, Gustave Roussy, Villejuif, France; Institut National de la Santé et de la Recherche Médicale – UMR935/UA9, Université Paris-Saclay, Villejuif, France; INGESTEM National IPSC Infrastructure, Université de Paris-Saclay, Villejuif, France; Université de Paris, Faculté des Sciences Pharmaceutiques et Biologiques, Paris, France; Centre Léon Bérard, Lyon, France; Université Claude Bernard, Lyon, France; Unicancer, Paris, France; Institut National de la Santé et de la Recherche Médicale, U981, Gustave Roussy, Villejuif, France; INSERM UMR 976, Institut de Recherche Saint-Louis, Université de Paris, Paris, France; Laboratory of Immunology, AP-HP, Hôpital Saint Louis, INSERM UMR1149, Université de Paris, Paris, France; Institut Hospitalo-Universitaire Méditerranée Infection, Marseille, France; Centre de Recherche des Cordeliers, Equipe labellisée par la Ligue contre le cancer, Université de Paris, Sorbonne Université, Inserm U1138, Institut Universitaire de France, Paris, France; Metabolomics and Cell Biology Platforms, Gustave Roussy Cancer Center, Université Paris Saclay, Villejuif, France; Pôle de Biologie, Hôpital Européen George Pompidou, Assistance Publique – Hôpitaux de Paris, Paris, France; Université de Paris, Paris, France; Department of Women’s and Children’s Health, Karolinska Institute, Karolinska University Hospital, Stockholm, Sweden; Suzhou Institute for Systems Biology, Chinese Academy of Medical Sciences, Suzhou, China; Medizinische Klinik, Johannes Gutenberg University Mainz, Germany; Division of Infection and Immunity, University College London, London, UK; NIHR Biomedical Research Centre, University College London Hospitals, London, UK

**Keywords:** TH2/TH1 paradigm, cancer, COVID-19, S1-RBD, viral variants, T cell memory

## Abstract

Optimal vaccination and immunotherapy against coronavirus disease COVID-19 relies on the in-depth comprehension of immune responses determining the individual susceptibility to be infected by SARS-CoV-2 and to develop severe disease. We characterized the polarity and specificity of circulating SARS-CoV-2-specific T cell responses against whole virus lysates or 186 unique peptides derived from the SARS-CoV-2 or SARS-CoV-1 ORFeome on 296 cancer-bearing and 86 cancer-free individuals who were either from the pre-COVID-19 era (67 individuals) or contemporary COVID-19-free (237 individuals) or who developed COVID-19 (78 individuals) in 2020/21. The ratio between the prototypic T helper 1 (TH1) cytokine, interleukin-2, and the prototypic T helper 2 (TH2) cytokine, interleukin-5 (IL-5), released from SARS-CoV-2-specific memory T cells measured in early 2020, among SARS-CoV-2-negative persons, was associated with the susceptibility of these individuals to develop PCR-detectable SARS-CoV-2 infection in late 2020 or 2021. Of note, T cells from individuals who recovered after SARS-CoV-2 re-infection spontaneously produced elevated levels of IL-5 and secreted the immunosuppressive TH2 cytokine interleukin-10 in response to SARS-CoV-2 lysate, suggesting that TH2 responses to SARS-CoV-2 are inadequate. Moreover, individuals susceptible to SARS-CoV-2 infection exhibited a deficit in the TH1 peptide repertoire affecting the highly mutated receptor binding domain (RBD) amino acids (331-525) of the spike protein. Finally, current vaccines successfully triggered anti-RBD specific TH1 responses in 88% healthy subjects that were negative prior to immunization. These findings indicate that COVID-19 protection relies on TH1 cell immunity against SARS-CoV-2 S1-RBD which in turn likely drives the phylogenetic escape of the virus. The next generation of COVID-19 vaccines should elicit high-avidity TH1 (rather than TH2)-like T cell responses against the RBD domain of current and emerging viral variants.

## Introduction

The emergence and spread of severe acute respiratory syndrome coronavirus-2 (SARS-CoV-2), the causative agent of coronavirus disease 2019, have resulted in devastating morbidities and socioeconomic disruption. The development of community protective immunity relies on long-term B and T cell memory responses to SARS-CoV-2. This can be achieved through viral infection [1] or by vaccination [2–4]. Reports on rapidly decreasing spike- and nucleocapsid-specific antibody titers post-COVID-19 infection [5] or reduced neutralizing capacity of vaccine-induced antibodies against viral escape variants compared to the ancestral SARS-CoV-2 strain [6, 7] have shed doubts on the importance of humoral immunity as a standalone response. In contrast, T cell immunity was identified as an important determinant of recovery and long-term protection against SARS-CoV-1, even 17 years after infection [8–11].

The TH1 versus TH2 concept suggests that modulation of the relative contribution of TH1 or TH2 cytokines regulates the balance between immune protection against microbes and immunopathology [12–14]. TH1 cells (as well as cytotoxic T cells with a similar cytokine pattern referred to as TC1 cells) produce IFNγ, IL-2, and TNFβ, promote macrophage activation, antibody-dependent cell cytotoxicity, delayed type hypersensitivity, and opsonizing and complement-fixing IgG2a antibody production [12]. Therefore, TH1/TC1 cells drive the phagocyte-dependent host response and are pivotal for antiviral responses [13, 14]. In contrast, TH2 (and TC2) cells produce IL-4, IL-5, IL-6, IL-9, IL-10, and IL-13, providing optimal help for both humoral responses and mucosal immunity, through the production of mast cell and eosinophil growth and differentiation factors, thus contributing to antiparasitic and allergic reactions. Naïve T cell differentiation to distinct TH fates is guided by inputs integrated from TCR affinity, CD25 expression, costimulatory molecules, and innate cell–derived cytokines [15].

SARS-CoV-2-specific T cell immunity plays a key role during acute COVID-19, and up to eight months after convalescence [16–20]. Indeed, functional T cell responses remain increased in both frequency and intensity up to six months post-infection [5]. They are mainly directed against spike, membrane and nucleocapsid proteins, and have been studied in greater detail by single cell sequencing in a limited number of patients. Memory TH1/TC1 T cells specific for SARS-CoV-2 and follicular T helper cells (TFH) cells have been detected in mild cases [21]. However, cases of reinfection have been reported [22] raising questions on the clinical significance of T cell polarization and peptide repertoire specificities against current viral variants. Moreover, pioneering reports suggest that, before SARS-CoV-2 became prevalent (i.e., before 2020), some individuals exhibit immune responses, mainly among CD4^+^ T cells, against SARS-CoV-1 nucleocapsid (NC) and ORF1a/b, or common cold coronaviruses (CCC) spike and nucleocapsid proteins that are cross-reactive with SARS-CoV-2 [9, 23, 24, 80]. However, the relevance of CCC or SARS-CoV-1-specific memory T cells for effective protection against the current pandemic remains questionable [21, 25].

In the present report, we studied SARS-CoV-2 –specific T cell responses in 382 cancer-bearing or cancer-free subjects, and prospectively followed up 227 COVID-free individuals to understand which T cell polarity and peptide repertoire may convey resistance to COVID-19. We found that a SARS-CoV-2-specific IL-2/IL-5 lymphokine ratio<1 conferred susceptibility to COVID-19 infection or re-infection, coinciding with defective TH1 recognition of the receptor binding domain (RBD) of the spike protein, likely affecting viral evolution by selecting for new antigenic variants.

## Results

### Effector and memory T cell responses against coronaviruses during COVID-19 infection

We conducted a cross-sectional analysis of the functional T cell responses across several cohorts of healthy individuals and cancer patients enrolled in various prospective studies (Table S1-S2). First, an epidemiological observational study (Figure 1A) aimed at defining the incidence, prevalence, and severity of COVID-19 in cancer patients, opened during the first surge of the pandemic at Gustave Roussy [26]. While the majority of the 227 enrolled cancer patients stayed COVID-19-free, we could analyze T cell responses during the acute phase of SARS-COV-2 infection (n=24) or during convalescence post-COVID-19 (n=28). All the other “unexposed” (COVID-19-negative) cancer patients were analyzed anytime between mid-March and September 2020 (Table S1, Figure 1A). In parallel, we analyzed 22 cancer-free, COVID-19-free healthy volunteers (HV) from 15 distinct families at the same time as their family members were in convalescent phase for COVID-19 (n=26) (Figure 2A, Table S3). A third cohort of 67 individuals from the pre-COVID-19 era (leukocytes frozen between 1999 and 2018) were either HV from the blood bank (n=38) or cancer patients (n=29) recruited in the context of clinical trials [27–30] (Table S1, Figure 1A).

**Figure 1.**
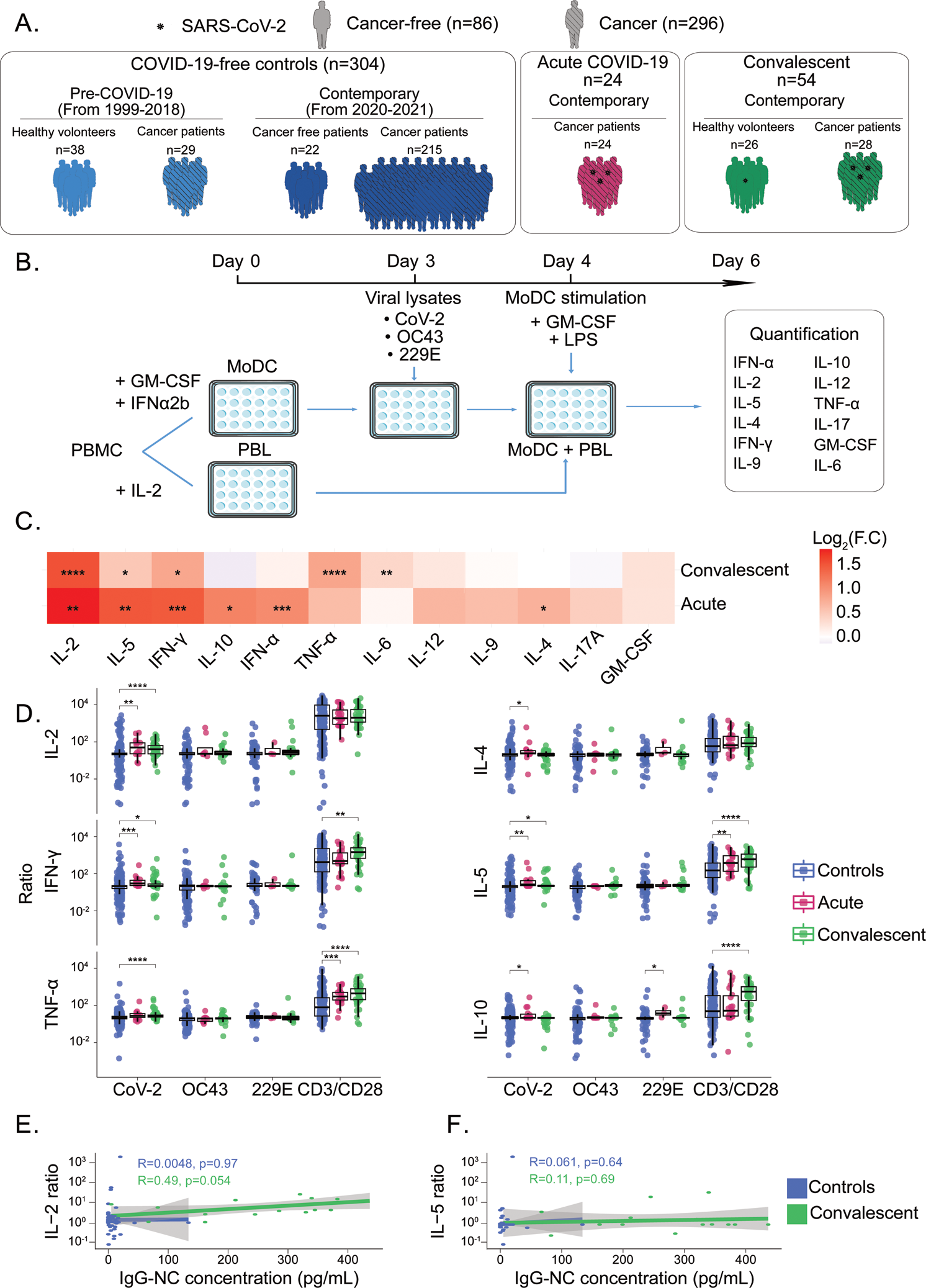
SARS-CoV-2 TH1/TC1 responses in COVID-19 and unexposed individuals. A. Graphical representation of the prospective patient cohorts used for the study (refer to Table S1 and Table S2). **B.** First experimental in vitro stimulation assay of peripheral blood lymphocytes (PBL) using cross-presentation of viral lysates by autologous dendritic cells (DC). Twelve plex flow cytometric assay to monitor cytokine release in replicates. **C.** Mean fold changes (Log2, F.C) between SARS-CoV-2-specific cytokine secretions of acute COVID-19 patients or convalescent COVID-19 individuals and controls (also refer to S1C). **D.** Ratios of cytokine secretion between PBL stimulated with DC pulsed with SARS-CoV-2 (or the other CCC lysates) *versus* VeroE6 (or versus CCC respective control cell lines), at the acute or convalescent phases of COVID-19. One typical example is outlined in Figure S1A. Each dot represents the mean of replicate wells for one patient (Controls, n=304; Convalescent COVID-19, n=54; Acute COVID-19, n=24). Asterisks indicate statistically significant differences in comparison to the control group determined using two-sided Wilcoxon-Mann-Whitney test (\**p*<0.05, \*\**p*<0.01, \*\*\**p*<0.001, \*\*\*\**p*<0.0001). **E.** Spearman correlations between SARS-CoV-2-specific IL-2 (left panel) or IL-5 (right panel) release and anti-NC IgG antibody titers (Controls, n=63; Convalescent, n =16).

**Figure 2.**
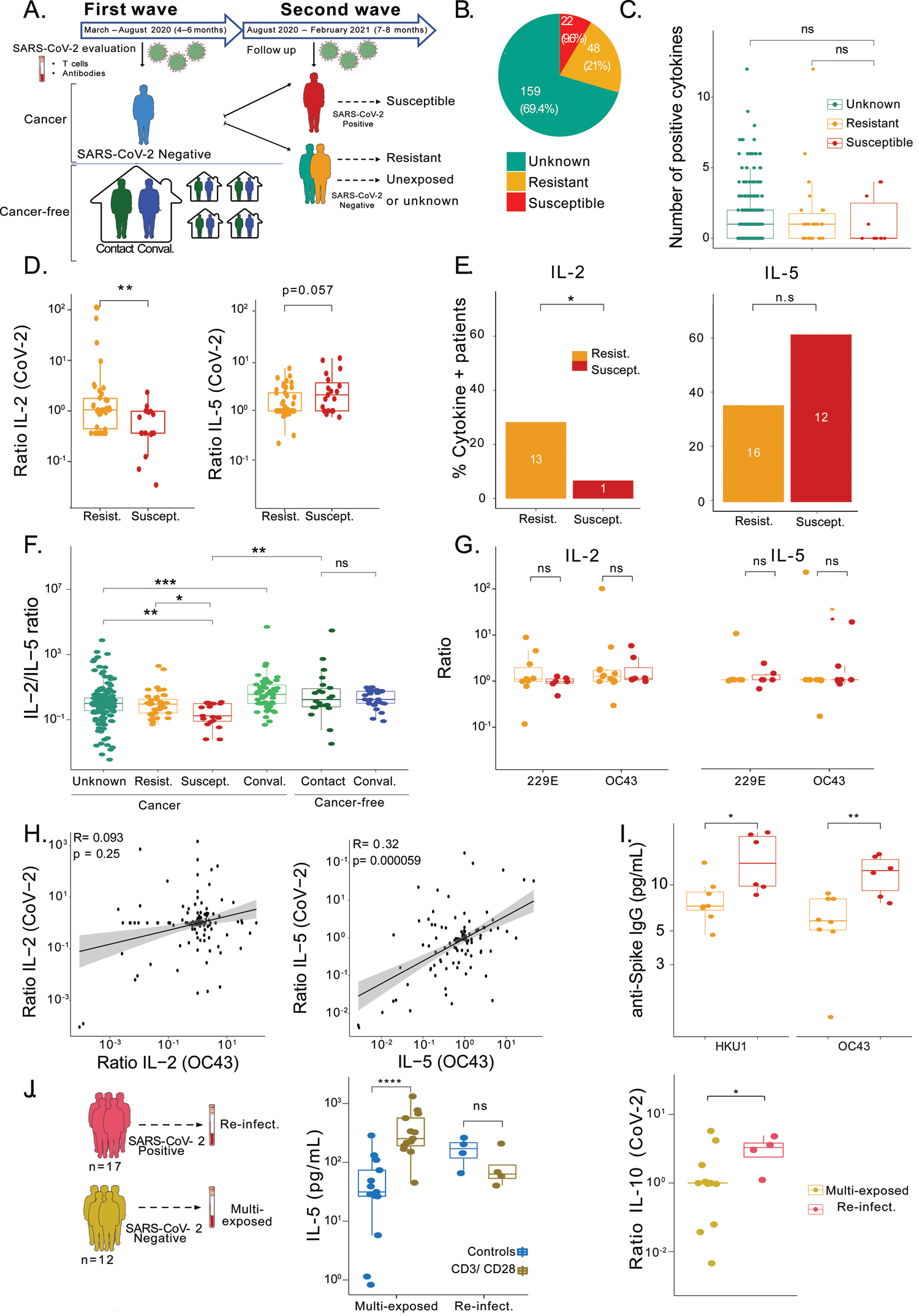
Unexposed individuals susceptible to COVID-19 exhibited a SARS-CoV-2 specific TH2 profile during the first surge of the pandemic. A-B. Upper scheme: Outline of the prospective collection of blood samples used to identify COVID-19 resistant (yellow) *versus* susceptible (red) cancer patients (A, upper panel, Table S1, Table S4) and pie chart indicating the absolute numbers (and %) of patients reported as contact (resistant) or infected (susceptible) or unexposed (green) during one-year follow-up (B). Lower scheme: Outline of the prospective collection of blood samples used for the comparison of T cell responses in the cohort of cancer-free individuals who lived in the same household with family members tested positive for COVID-19 during the 2020 lock down (Table S3). **C.** Number of positive cytokines released by SARS-CoV-2-specific PBL during the cross-presentation assay (Figure 1B) in each group (Unexposed, n=159; Resistant, n=48; Susceptible, n=22). **D-E.** SARS-CoV-2-specific IL-2 (left panel) and IL-5 (right panel) secretion contrasting resistant (yellow) *versus* infected (red) cases. Each dot represents the ratio (D) of the replicate wells in one individual and the box plots indicate medians, 25th and 75th percentiles for each patient subset. The bar plots (E) represent the percentage of positive patients (resist., n=41; suscept., n=19). Fisher exact test to compare the number of cytokine positive patients across groups (\**p*<0.05). **F.** SARS-CoV-2-specific IL-2/IL-5 ratios (means±SEM) in the different subsets presented in panel A. Refer to Figure S4 for the waterfall plots to visualize variations in the percentages of individuals with IL-2/IL-5 ratios > or < 1 according to subject category. **G**. CCC (OC43 and 229E)-specific IL-2 ratio (left panel) or IL-5 secretion ratio (right panel) contrasting contact (resistant, yellow dots, n=34) *versus* infected (susceptible, red dots, n=11) cases. **H.** Spearman correlations between OC43 *and* SARS-CoV-2-specific IL-2 (left panel) and IL-5 (right panel) secretions in 156 controls. **I.** Anti-spike IgG titers (means±SEM) specific of seasonal betacoronaviruses in contact (resist., yellow dots, n=34) *versus* infected (suscept., red dots, n=11) cases. **J**. Scheme detailing the two groups of cancer-free individuals from the same hospital with opposite clinical phenotypes (multi-exposed individuals (n=12) versus patients re-infected with SARS-CoV-2 (n=17) patients) (J, left panel). Results of the cross-presentation assay against SARS-CoV-2 for IL-10 (J, right panel) and IL-2 (refer to Figure S5B). IL-5 levels at baseline and after TCR cross-linking are depicted in the middle panel. Each dot represents the mean of two replicates for one patient. All group comparisons were performed using two-sided Wilcoxon-Mann-Whitney test and asterisks indicate statistically significant differences (\**p*<0.05, \*\**p*<0.01, \*\*\**p*<0.001, \*\*\*\**p*<0.0001).

T cell responses directed against viral lysates from the historic SARS-CoV-2 strain IHUMI846 (CoV-2) isolated in early 2020 or two endemic common cold coronaviruses (CCC), OC43 and 229E, were evaluated by an *in vitro* stimulation assay (IVS). As outlined in Figure 1B, in a first step, autologous dendritic cells (DC) were differentiated from monocytes for 72hrs in GM-CSF+IFNα2b. DC were spinoculated with viral particles from heat-inactivated viral lysates and activated with LPS in a second step. Peripheral blood lymphocytes (PBL) kept in low-dose IL-2 during this first step were then stimulated for 48hrs by activated DC after removal of LPS (Figure 1B). The specific viral lysates were compared to supernatants from cell lines permissive for viral replication (such as Vero E6 for SARS-CoV-2, HCT8 for OC43, MRC5 for 229E). Negative controls were unloaded DC/PBL cell cocultures, while positive controls were PBL stimulated with anti-CD3/anti-CD28-coated beads. The cytokines accumulating in the supernatants were analyzed by means of a 12-plex flow cytometry-based bead assay (Figure S1A). In this cross-presentation assay, SARS-CoV-2-related IFNγ or IL-2 secretions from PBL depended on MHC class I and MHC class II molecules, as shown using specific neutralizing antibodies (Figure S1B). We calculated the ratio of cytokine release by dividing interleukin concentrations following exposure to viral lysates by those obtained with the respective control supernatants, to ascribe the specificity of the reactivity to SARS-CoV-2 or to CCC antigens for each subject.

First, we characterized the intensity and the quality of PBL responses elicited at the acute phase of SARS-CoV-2 infection (day of symptom onset and/or first positive qPCR of the oropharyngeal swab and/or serology), between mid-March and mid-May 2020 in 24 interpretable tests performed on COVID-19^+^ subjects compared to a cohort of 304 controls Table S2). Robust SARS-CoV-2 specific IL-2 and IFNγ release, most likely caused by TH1/TC1 cells, and the secretion of IL-4, IL-5 and IL-10, most likely mediated by TH2/TC2 effector T cells, were detectable, and were dominated by IL-2 and IL-5, respectively (Figure 1C, Figure 1D). An antiviral response, characterized by type 1 IFN release, was also prevalent (Figure S1C), as previously detected by single cell transcriptomics in the subset of transitional memory CD4^+^ T cells [21]. Of note, COVID-19 infection did not reactivate CCC-specific T cell responses (Figure 1D, Figure S2A). Interestingly, the systemic T cell tone was shifted towards an inflammatory TH2 profile during the acute phase of the infection, as suggested by increased IL-5, IL-6, TNFα, and IL-17 secretion following TCR cross-linking in COVID-19 patients compared with individuals without COVID-19 (Figure 1D, Figure S1C).

We next examined the polarization of SARS-CoV-2-specific memory T cell responses between May and September 2020 in 54 convalescent COVID-19 individuals (median time lapse between PCR negative and T cell assay: 85 days, range: 13-106 days) compared with contemporary controls (Figure 1A, Figure 1C, Figure 1D, Figure S2, Table S2). A dominant SARS-CoV-2-specific memory TH1 response (resulting in the secretion of IFNγ, IL-2 and TNFα) prevailed in most convalescent subjects within the next 2-3 months after acute infection (Figure 1D). Of note, SARS-CoV-2 specific IL-2 release at recovery correlated with an increase in the frequency of circulating non-activated TFH cells (Figure S1D). In addition, SARS-CoV-2 specific IL-2 secretion was the only parameter correlating with anti-SARS-CoV-2 nucleocapsid (NC) antibody titers (reported to be stable for 8 months [5]) but not with IgG and IgA antibodies targeting the S1 domain of the SARS-CoV-2 spike protein including the RBD (Figure 1E-F and not shown).

More than 30% convalescent COVID-19 individuals displayed polyfunctional T cell responses defined by at least 2 cytokines with a secretion ratio >1.5 (Figure S2A-B). Memory TH1 (IL-2, IFNγ) responses were comparable in both healthy and cancer subsets of COVID-19 positive individuals (Figure S2B-C). As previously described [21, 23, 24, 31, 32], contemporary COVID-19 negative subjects also harbored significant SARS-CoV-2 specific-TH1/TC1 memory responses that appear to pre-exist in cancer patients and healthy individuals whose blood was drawn in the pre-COVID-19 era, including prior to outbreaks of SARS-CoV-1 and MERS (Figure S2B-C). The unsupervised hierarchical clustering considering 12 cytokines monitored in 358 subjects did not segregate pre-COVID-19 from contemporary unexposed individuals nor convalescent patients (Figure S2A).

Hence, while polyfunctional T cell responses dominated by IL-2 and IL-5 were elicited during the acute phase of COVID-19, after convalescence, recirculating memory T cells exhibited a TH1/TC1 profile in about 70% of COVID-19 patients, correlating with anti-SARS-CoV-2 NC antibody titers.

### Clinical relevance of the IL-2/IL-5 ratio to predict COVID-19 infection

We next determined the clinical significance of these memory T cell responses monitored in unexposed individuals during the first surge of COVID-19 (mid-March to mid-May 2020) to decipher the nature of memory T cells contributing to susceptibility or resistance to the successive surges of this viral pandemic in fall 2020 and winter 2021. We phoned 229 patients to discover that 22 individuals had developed COVID-19 infections (diagnosed by qPCR or serology) with different degrees of severity according to WHO criteria (Table S1, Figure 2A, Table S4). Indeed, about one third of the initially COVID-19-free individuals became contact cases (n=70) and 29% among these contact cases were diagnosed with COVID-19 infection by specific RT-qPCR or serology (n= 22, Figure 2B, Table S4). The unsupervised hierarchical clustering of the T cell secretory profiles in these 70 individuals failed to correctly segregate resistant (contact) from susceptible (infected) cases (Figure S3A). In addition, the polyfunctionality of T cell responses failed to segregate the two categories of individuals (Figure 2C, Figure S4B). However, two lymphokines, IL-2 and IL-5, secreted by memory T cells responding to SARS-CoV-2 correlated with resistance and susceptibility to SARS-CoV-2, respectively (Figure 2D-E). Indeed, the levels of IL-2 in the recall response and the proportions of individuals exhibiting IL-2 polarized T cell memory responses were both associated with resistance to COVID-19 (Figure 2D, *p*=0.01, two-sided Wilcoxon-Mann-Whitney test, Figure 2E, *p*=0.049, Fisher exact test). In contrast, IL-5 levels in recall responses were associated with increased susceptibility to COVID-19 (Figure 2D, *p*=0.057, two-sided Wilcoxon-Mann-Whitney test). Resistant individuals harbored a coordinated SARS-CoV-2 specific-TH1 response (IFNγ, TNFα) in mid-March to mid-May 2020, while subjects susceptible to COVID-19 exhibited inflammatory TH9/TH17-like networks (Figure S4C-D). These findings are reminiscent of data segregating asymptomatic or mild from severe diseases [33, 34]. For this reason, we henceforth analyzed the clinical significance of the ratio between SARS-CoV2-specific IL-2 and IL-5 release. Indeed, the IL-2/IL-5 recall response ratio was significantly higher in cancer patients who were SARS-CoV-2-resistant (Figure 2F) and in convalescent patients (Figure S4A). The vast majority (16 out of 19) of cancer patients doomed to be infected with SARS-CoV-2 harbored an IL-2/IL-5 ratio ≤1, with the two severe COVID-19 cases exhibiting an IL-2/IL-5 ratio <10 (Figure S4B). In contrast, CCC-specific T cell reactivities did not allow to discriminate susceptible from resistant individuals (Figure 2G), although IL-5 stood out as the strongest correlate between SARS-CoV-2 and CCC-specific T cell responses among 156 individuals with no correlation between SARS-CoV-2 and OC43-specific IL-2 secretion (Figure 2H). Reinforcing this notion, titers of IgG antibodies directed against the spike of the seasonal betacoronaviruses OC43 and HKU1 (but not the alphacoronavirus 229E and NL63) were higher in individuals susceptible to SARS-CoV2 compared to resistant individuals (Figure 2I and not shown).

The SARS-CoV2-specific IL-2/IL-5 recall response ratio was also clinically significant in the cohort of cancer-free individuals that were locked down together with their COVID-19-positive family members (Figure 2A, Table S3, Figure S4D). Individuals who did not get infected harbored IL-2/IL-5 ratios>1 reaching mean values comparable to those achieved in convalescent individuals (Figure 2F, Figure S4A) at higher frequencies than the overall population (Figure S4C).

We next compared the immunogenicity of the lysates derived from the original SARS-CoV-2 strain (IHUMI846) with that of the Danish (IHUMI2096, 20A.EU2, B.1.367, GH) and North African (IHUMI2514, 20C, B.1.160, GH) strains isolated at the end of 2020 [35]. Of note, T cells lost their capacity to produce IL-2 in response to the IHUMI2096 and IHUMI2514 viral variants while IL-17 release tended to increase (p= 0.0857, Figure S5A).

Given that 15-20% of convalescent individuals mounted a TH2/TC2 memory response to SARS-CoV-2 (Figure S2A), we wondered whether these individuals could be at higher risk to get re-infected by SARS-CoV-2 variants. Hence, we analyzed PBMCs in a series of individuals (n=12) who were diagnosed with COVID-19 during the first surge of the SARS-CoV-2 pandemic and then were re-infected with a viral variant prevailing during the later outbreak occurring in fall 2020 or winter 2021, comparing them to health care workers who were multi-exposed cases with multiple negative oropharyngeal SARS-CoV-2-specific PCR assays (n=17) (Table S5, Figure 2J, left panel). Surprisingly, monocytes could be differentiated into dendritic cells only in 4 re-infected patients. Unstimulated PBL from re-infected and re-convalescent subjects spontaneously secreted much higher levels of IL-5 than did PBL from multi-exposed cases, reaching similar ranges as those obtained after TCR crosslinking (Figure 2J, middle panel). After specific re-stimulation with SARS-CoV-2 lysates, IL-10 was markedly increased in recall responses from re-infected but not multi-exposed cases (Figure 2J, right panel) and IL-2 was undetectable (Figure S5B). In this small cohort of multi-exposed individuals, the recognition pattern of the United Kingdom (UK) (IHUMI3076, B.1.1.7), South Africa (IHUMI3147, B.1.351) and Brazil (IHUMI3191, P.1) [35] strains were quite variable, some individuals losing the TH1/TC1 or acquiring a TH2/TC2 profile, depending on the strain (Figure S5C).

We conclude that the TH1/TC1 *versus* TH2/TC2 polarity of SARS-CoV-2 specific-memory T cell responses is associated with effective clinical protection against infection, an IL-2/IL-5 ratio >1 indicating resistance to the COVID-19.

### Defects in the TH1/TC1 response against the SARS-CoV-2 RBD of spike glycoprotein in susceptible individuals

In hosts affected by viral infections or cancer, the breadth of T cell epitope recognition is a prerequisite for protective immunity [36–38]. We analyzed the diversity of SARS-CoV-2 T cell responses by single peptide mapping using 186 peptides with 9 to 51 amino acids corresponding to 146 non-overlapping or poorly overlapping epitopes of the SARS-CoV-2 ORFeome (among which 25 epitopes were shared with SARS-CoV-1), encompassing membrane, nucleocapsid, spike, ORF3a, ORF8 and ORF10 proteins, plus 41 epitopes covering the SARS-CoV-1 ORFeome of immunological relevance (among which 8 epitopes were shared with SARS-CoV-2), as well as a series of positive controls, namely epitopes from Influenza virus, Epstein Barr virus (EBV), Cytomegalovirus (CMV), phytohemagglutinin (PHA), and anti-CD3ɛ (OKT3) antibody. IFNγ responses against these 186 peptides were evaluated in 164 individuals (123 unexposed individuals, 25 convalescent COVID-19-positive patients, 16 re-infected patients, see Table S6). To enable the detection of low-frequency SARS-CoV-2 peptide-specific T cells, we used an *in vitro* 7 day-long, IL-2+IL-15 enriched IVS assay in the presence of each individual peptide in duplicates (Figure 3A). We chose to monitor IFNγ, a proxy for TH1/TC1 responses, as opposed to IL-2, in the 7 day-coculture supernatants by ELISA because recombinant human IL-2 was already added to the IVS assay (Figure 3A). The overall recognition patterns of these peptides across various patient populations, and their individual frequencies are detailed in Figure 3B, and Figure S6-S7. About 10% convalescent individuals recognized more than 15% of our peptide selection within the SARS-CoV-2 ORFeome (Figure S6 inset). T cell responses in unexposed patients, in particular in the pre-COVID-19 era, covered large specificities, as suggested by previous reports [9, 21, 24] (Figure 3C, right panel and Figure S6). In accord with the literature [9, 24], the immune response of convalescent COVID-19 patients was mainly directed against spike, membrane, and nucleocapsid (NC) and, to a lower extent, against ORF3a, ORF8, and ORF10 (Figure 3B, 3D left panel). The breadth of the peptide recognition coverage was not significantly reduced in cancer patients compared with others (Figure 3B, 3C left panel). In a limited number of individuals, we measured not only IFNγ but also IL-5, IL-9 and IL-17 by ELISA. The recognition profile specific to the spike (and more specifically the RBD) as well as ORF8 was more geared toward TH1/TC1 (IFNγ) than TH9 or TH2 (IL-9 or IL-5 production respectively) (Figure S8A-C). The membrane- and NC-specific repertoire was strongly TH17 oriented (Figure S8B).

**Figure 3.**
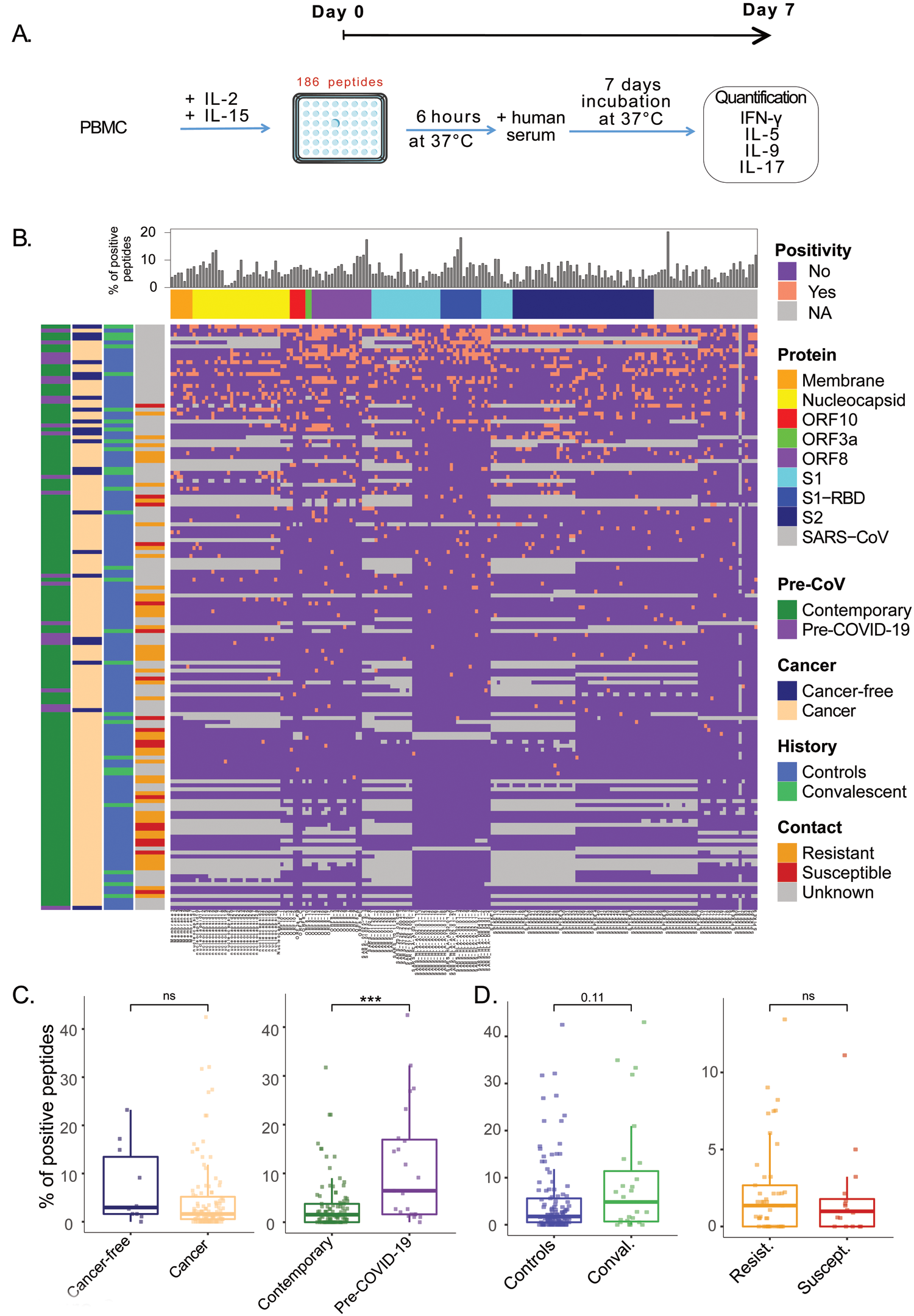
Peptide recognition patterns in all distinct subsets of individuals: repertoire breadth of peptide does not predict resistance to COVID-19. A. Experimental setting for the 186 peptide-based *in vitro* stimulation assays. **B.** Bicolor map of peptide recognition (positive in salmon, negative in purple, not determined in grey). Patients (n=164) were ordered in columns by unsupervised hierarchical clustering and peptides were ordered on rows according to recognition frequency. The left column details clinical information and the upper line indicates the peptide frequency and the names of the proteins. **C-D.** Percentages of positive peptides in individuals from the pre-COVID19 era (n=23) *versus* contemporary controls (n=100) (C, right panel) and in cancer (n=112) *versus* cancer free contemporary individuals (n=13) (C, left panel) and in uninfected (control, n=123) *versus* convalescent (recovery, n=25) (D, left panel) and resistant individuals (non-infected contact cases (n=45) *versus* susceptible (infected, n=18) individuals (D, right panel). Group comparisons were performed using two-sided Wilcoxon-Mann-Whitney test and asterisks indicate statistically significant differences (\**p*<0.05, \*\**p*<0.01, \*\*\**p*<0.001, \*\*\*\**p*<0.0001).

Using logistic regression analyses, we determined the TH1/TC1 peptide recognition fingerprint significantly associated with each patient category (Figure S9). The hallmark repertoire of the pre-COVID-19 era consisted in a stretch of peptides covering part of the SARS-CoV-1 genome (spike, membrane, ORF3a, NC), some peptide residues sharing high or complete homology with SARS-CoV-2, as well as numerous ORF8 sequences (Table S6). Of note, the recognition pattern of these SARS-CoV-1 epitopes highly correlated with responses directed against ORF8 peptides (not shown). In contrast, the COVID-19-associated blueprint encompassed many nucleocapsid peptides (NC_1 (residues 1-15), NC_6-7 (residues 76-105, NC_8 (residues 106-120) sharing 93% and 100% homology with OC43 and HKU1, respectively, the HLA-A2-restricted nonamer (RLNQLESK<u>V</u>) NC_226-234 from SARS-CoV-1 (sharing high structural homology with the SARS-CoV2 epitope RLNQLESK<u>M</u>) and another SARS-CoV-1 NC nonamer peptide (NC_345-361), three peptides residing in ORF8, two epitopes belonging to the RBD region (“SPIKE29”) found at high frequency across subjects (23.5%), as well as a peptide from the C terminal portion of spike (“SPIKE84”, residues 1246-1260) (Figure S9A). Cancer patients tended to lack some specificities, yet with no prototypical signature (Figure S9A).

Next, we investigated the ORFeome peptide repertoire associated with SARS-CoV-2-specific IL-2 memory responses in 118 unexposed individuals by means of linear regression analysis (Figure 4A, left panel). Among the 9 peptides associated with a positive contribution to IL-2 secretion, one nonamer (KLPDDF<u>M</u>GCV in SARS-CoV-1 genome and KLPDDF<u>T</u>GCV in the SARS-CoV-2 genome) resided in the RBD region that constitutes the binding site for its cellular receptor angiotensin-converting enzyme 2 (ACE2) [39] while, conversely among the 13 peptides associated with a hole in the TH1 response, 5 resided within the RBD of the spike glycoprotein. More specifically, there was a statistically significant enrichment of RBD-related peptides within this TH1/TC1 hole (Figure 4A, right panel).

**Figure 4.**
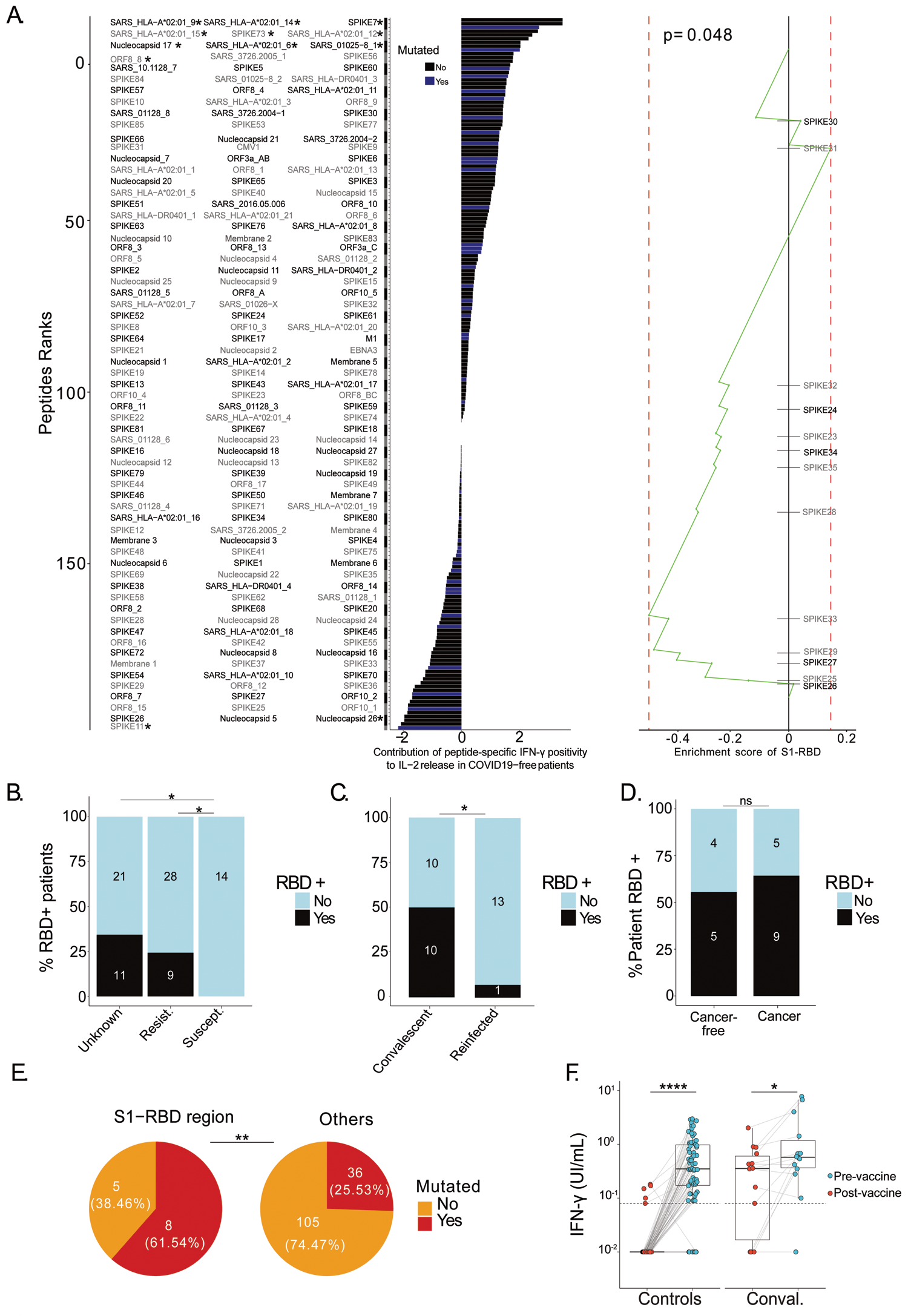
Receptor Binding Domain (RBD)-directed TH1/TC1 recall responses predict resistance to COVID-19. A. Linear regression analysis of the relative contribution (t-value corresponding to the regression coefficient) of each peptide to SARS-CoV-2-specific TH1/TC1 responses (measured as IL-2 secretion in responses to whole virus lysate in Fig. 1), as determined in the peptide-specific IFNγ secretion assay in 123 COVID-19 negative individuals. Statistically significant peptides (*p*<0.05) are annotated with asterisks (A, left panel). Peptides colored in blue reportedly harbor at least one mutation within SARS-CoV-2 variants (Table S7). Peptide set enrichment analysis plot (A, right panel). The contribution of each peptide to the SARS-CoV-2-specific IL-2 secretion was used to rank 164 peptides. The enrichment score of S1-RBD peptides suggested that this peptide set presented lower t-values than randomly expected (*p*=0.048) (A, right panel). **B-C**. Percentages of patients recognizing at least one S1-RBD peptide in the IFNγ ELISA of the peptide IVS assay across patients’ groups described in Figure 2A (B) or convalescent versus re-infected patients (C). **D.** Percentages of patients recognizing at least one S1-RBD peptide in the IFNγ ELISA of the peptide IVS assay in cancer and cancer free patients from the pre-COVID19 era. **E.** Percentages and absolute numbers of mutations contained in our S1-RBD peptide list reported in the current SARS-CoV-2 variants (refer to Table S7). The difference of the probability of mutation in S1-RBD region and in other regions was evaluated using logistic regression (Odd Ratio=0.21, 95% confidence interval [0.06; 0.68], *p*=0.01). **F.** IFNγ levels in whole blood samples from a cohort of COVID-19 negative or positive (convalescent) HCW (refer to Table S10) drawn pre- and post-vaccines, that were measured after a 22-hour stimulation with mitogens using a pool of overlapping RBD peptides (BioMérieux assay). Group comparisons were performed using two-sided paired Wilcoxon-Mann-Whitney test and asterisks indicate statistically significant differences (\**p*<0.05, \*\**p*<0.01, \*\*\**p*<0.001, \*\*\*\**p*<0.0001).

In order to validate the clinical significance of the TH1/TC1 repertoire hole and the assumption that a defect in the TH1/TC1 recognition pattern of the RBD sequence could be a risk factor for COVID-19, we calculated the number of positive peptides selected from the RBD region spanning aminoacid 331-525 residues (called “SPIKE23” to “SPIKE35” in Table S6), in each of the 83 individuals who were comprehensively explored in the peptide-based IVS assay, 37 resistant (contact) individuals, 14 infected persons (susceptible) as well as 32 controls (unexposed lockdown and/or unknown) using the IFNγ ELISA (Table S1). While susceptible individuals exhibited a significant defect in the RBD-related TH1/TC1 repertoire (Figure 4B), up to 25% of the resistant individuals harbored robust TH1/TC1 responses to the 331-525 aminoacid residues of RBD (Figure 4B, *p*=0.049, Fisher exact test). Again, the RBD-specific TH1/TC1 responses were almost undetectable in patients who got infected twice with SARS-CoV-2 (Table S5), while they could be measured in 50% convalescent COVID-19 patients (Figure 4C, *p*=0.011, Fisher exact test) in accordance with a recent report highlighting the immunodominance of the S346-365 region (corresponding to our “SPIKE24” epitope) in convalescent individuals [40]. Such S1-RBD-specific TH1 responses pre-existed in the pre-COVID-19 era at the same frequency in cancer or cancer-free individuals (Figure 4D). T cell responses directed against S1-RBD peptides were evaluated for both IL-5 and IFNγ secretions in 36 patients. They almost exclusively exhibited a TH1 pattern (in 10/36 cases), with only 3/36 individuals harboring a TH2 profile (Table S7). Of note, there was a robust consistency and concordance of the polarization status of patients between the two (cross-priming and peptide-based) IVS assays (McNemar test, p= p.val=2,2e-16 comparing SARS-CoV-2 or peptide-specific IL-2 and IFNγ release, McNemar test, p= p.val=2.2e-16 comparing SARS-CoV-2 or peptide-specific IFNγ release, p.val <1e-16).

We performed immunoinformatic-based prediction analyses of S1-RBD peptide binding affinities to MHC class I and class II molecules using the NetMHCpan algorithm. This approach revealed strong binding affinities to MHC class I HLA-A, -B and -C alleles for the RBD epitopes “SPIKE25” (residues 361_375), “SPIKE27” (residues 391-405), “SPIKE31” (residues 451-465). In contrast, “SPIKE33” (residues 481_495) was predicted to have a low or absent affinity for HLA-B and HLA-C alleles, respectively (Table S8a). Only “SPIKE24”, “SPIKE25” and “SPIKE31 were predicted to bind with a high affinity to HLA-DR alleles (Table S8b) as already reported for the immunodominant S346-365 region [40].

In accordance with the immunodominance of S1-RBD, the other signatures indicated by our logistic regression analysis (Figure S9A), namely the convalescent or the pre-COVID-19 era-related blueprints were not significantly associated with COVID-19 resistance (Figure S9B-C).

Given that immunoselection may drive antigenic drift of viruses as well as the evolution of viral phylogeny, we analyzed the coincidence of hot spot mutations in the SARS-CoV-2 ORFeome [41] with T cell memory patterns of clinical significance (Table S9). Of note, significantly higher mutation frequencies were detected within the S1-RBD-specific TH1 response (62%) compared with other regions of the SARS-CoV2 orfeome (26%) (Odd Ratio = 0.21, 95% confidence interval [0.06; 0.68], *p*=0.01, Figure 4E).

During the course of this study, two SARS-CoV-2 mRNA vaccines were approved by FDA and EMA based on reports that they prevent COVID-19 infection with an efficacy of >90% [3]. Using a simple whole blood stimulation assay allowing the detection of IFNγ-producing anti-viral T cells following peptide stimulation within 18 hours [42], we analyzed RBD-specific T cell reactivities before and after dual vaccination with BNT162b2 mRNA (BioNTech/Pfizer) in 70 unexposed health care workers (HCW) and 14 COVID-19 convalescent HCW (Table S10). The vast majority (approximately 90%) of naive vaccinees mounted robust RBD-specific TH1/TC1 cell responses after 2 immunizations (Figure 4F).

Hence, our findings suggest that defects in the TH1 repertoire affecting the recognition of SARS-CoV-2 S1-RBD are associated with susceptibility to infection or re-infection by SARS-CoV-2. This antigenic region appears to mutate more than other regions of the SARS-CoV-2 orfeome, perhaps as a result of an immune system-mediated evolutionary pressure.

## Discussion

Here, we unravel the first *prospective* correlation between preexisting (before the first surge) SARS-CoV-2-specific TH2/TC2 immune responses and susceptibility to infection with SARS-CoV-2 or re-infection with viral variants. Both in cancer patients and cancer-free subjects, the best immunological correlate with future SARS-CoV-2 infection was undistinguishably a recall response characterized by a low ratio of TH1/TH2 lymphokines (and more precisely an IL-2/IL-5 ratio <1) secreted upon exposure to the original SARS-CoV-2 viral strain. This recall response coincided with a hole within the TH1/TC1 cell repertoire affecting the RBD of the spike protein. Moreover, in a small series of re-infected individuals, maladaptive anti-SARS-CoV-2 responses were characterized by elevated baseline TH2 responses (spontaneous IL-5 release from PBL) and IL-10-centered recall responses. Finally, vaccinees immunized with a clinically approved mRNA encoding the spike protein mounted a robust RBD-specific TH1/TC1 response, that may account for immune protection.

Reportedly, CD4^+^ TH1 and TH2 responses are induced during the primary phase of viral infection, and both TH1 and TH2 can generate an anamnestic response upon rechallenge with the same virus [43]. Survivors from SARS-CoV-1 infection developed polyfunctional T cells producing TH1 cytokines and long-term CD8^+^ T cell responses as late as 11 years post-infection [9]. The TH1 cytokine IL-2 (which correlated with circulating non-activated TFH cells in convalescent patients in our study) was the pivotal factor allowing us to distinguish susceptible from resistant individuals. Signaling via the high-affinity IL-2 receptor (which requires CD25/IL-2Rα expression) favors the generation of CXCR5^-^ T effector cells, and this is associated with TH1 responses sustained by the transcription factor TBX21. Moreover, the development of IFNγ producing effector memory T cells depends upon CD25 [15]. Accordingly, upon infection with lymphocytic choriomeningitis (LCMV), CD25-deficient CD4^+^ T cells largely fail to form IFNγ producing T effector cells in secondary lymphoid organs and to generate lung tissue resident memory T cells [44].

In contrast, increased TH2 cytokine release correlated with poor outcome in patients, a finding corroborated in mouse studies of SARS-CoV-1 [45, 46]. During SARS-CoV-2 infection, TH2-associated blood markers, such as eosinophilia and circulating IL-5, IL-33, eotaxin-2 and eotaxin-3 correlated with COVID-19 severity, contrasting with the fact that CD4^+^ and CD8^+^ T cells from COVID-19 patients secrete comparable amounts of IFNγ in moderate and severe disease [47].

What cellular cues may influence TH2 differentiation or maintenance? Distinct dendritic cell subsets could specifically induce TH2 immunity, while limiting TFH responses and humoral immunity [48–50]. Indeed, preferential trans-vasation of blood-born cDC2 (CD1c^+^ DC) into the bronchoalveolar fluid was described for severe COVID-19 [51], and TH1/TH2 polarization is not only achieved during the priming response in lymphoid organs but is largely driven by the preferential expansion of either TH1 or TH2 cells in non-lymphoid tissues [52]. Various cell types located in peripheral tissues can impact T cell fate [53]. In the lungs of severe COVID-19 patients, a clonal expansion of IL-17A/F and GM-CSF producing TRM-like CD4^+^ T cells persisted even after viral clearance [34]. Our findings point to some correlative links between IL-5 and GM-CSF in recall responses from individuals susceptible to COVID-19. While IL-7 secreting stromal cells may promote survival of TH2 memory cells within lung tertiary lymphoid structures [54, 55], it remains unclear to which extent local (oropharyngeal or pulmonary) cues might influence the maintenance of SARS-CoV-2 specific or cross-reactive TH2 responses.

TCR signaling plays a major role in CD4^+^ polarization and can vary according to the TCR affinity, the amount of peptide/MHC-II complexes perceived by a TCR, or the length of time a T cell spends proofreading peptide/MHC-II complexes [15]. Of note, the RBD-specific TH1/TC1 responses against regions 361_375 and 391_405 of spike exhibited robust binding capacities across all MHC class I alleles. Several authors reported cross-reactivities between CCC and SARS-CoV2 [9, 20, 23, 24, 31, 32, 56, 57]. However, such cross-reactive T cells may turn out to be harmful with respect to clinical correlations [58–63]. Indeed, according to one report [21], preexisting CCC-specific memory CD4^+^ T cells exhibit low TCR avidity in almost all unexposed individuals, and are strongly expanded in severe COVID-19 but not in mild cases. Moreover, CCC/SARS-CoV-2-cross-reactive T cell clones shared among convalescent and infected individuals harbored lower functional avidity than non-cross-reactive clones, suggesting antigenic imprinting of the TCR repertoire by previous exposure to CCC [25, 82]. Of note, these spike-specific cross-reactive CD4^+^ T cells might not only re-expand during infection but also following vaccination. In line with this possibility, we detected a strong positive correlation between CCC and SARS-CoV2-specific IL-5 release by memory T cells in all unexposed individuals. Moreover, CCC-specific IgG titers were higher in susceptible compared to resistant individuals. Finally, the SARS-CoV-1 and ORF8-specific T cell repertoire prevailing in the pre-COVID-19 era failed to be clinically relevant for the avoidance of COVID-19 and such a repertoire was frequently detected in re-infected individuals during their convalescence phase. Hence, we cannot rule out the possibility that a preexisting TH2 immunity, for instance directed against sequences shared by sarbecoviruses [9] could increase the susceptibility to, and severity of, SARS-CoV-2 infection [45–47].

The landscape of prevalence and immunodominance of SARS-CoV-2 epitopes - supposedly associated with protection during the acute phase - has been thoroughly investigated [64]. Using 40-mer peptide pools covering regions of membrane, nucleocapsid, ORF3a, ORF7/8, and spike proteins, Tan et al. observed a statistically significant correlation between the early appearance of SARS-CoV-2 peptide-reactive cells and shorter duration of infection [65]. However, viruses employ numerous strategies to evade CD8^+^ T cell immune responses [66]. Our data obtained in COVID-19-free individuals who remained resistant to overt infection despite exposure to the virus strongly support the immunological and clinical relevance of memory TH1/TC1 responses directed against the spike S1-RBD region. An immune-driven selection process of viral phylogeny can occur [67]. Early (but transient) induction of ORF7/8-specific TH1 cell immune responses as well as antibody responses against ORF8 have already been reported during the acute phase of COVID-19 in small numbers of patients. Some arguments plead for the biological significance of ORF8-specific T cell immunity in viral control. Indeed, a 382-nucleotide deletion that truncates ORF7b and ORF8, culminating in the loss of ORF8 transcription, and conferring mild forms of infections [68] has been reported [69]. However, our data do not support the clinical relevance of ORF8-specific immune responses.

There is growing evidence of the links between mutations within the SARS-CoV-2 spike protein and the evasion of neutralizing antibody responses [70, 71]. Single nonsynonymous mutations in SARS-CoV-2 can theoretically subvert the immune response to CTL epitopes as well [72]. These studies suggested that immune selection may shape the mutational landscape of CD8^+^ T cell epitopes. Our data fuel the theory that i) robust TH1 memory immune responses against RBD might be important in restraining viral infection, thus exerting a selective pressure on the virus, obliging it to generate escape variants by mutation of RBD, ii) preexisting TH2 antiviral responses might not only be incapable of eliminating SARS-CoV-2-infected cells but actually favor (re-)infection with SARS-CoV-2, ultimately increasing the viral reservoir, thus favoring the emergence of viral variants.

Our work may have important consequences for the design of next-generation vaccines against COVID-19. Immunization strategies should aim at triggering SARS-CoV-2 specific TH1/TC1 (rather than TH2/TC2) responses. The efficacy of cellular immune response relies on three components, (i) the antigen, (ii) the adjuvant and (iii) the dynamics of viral evolution [73]. Immunization with inactivated SARS-CoV-1 or with the whole spike (S) protein, caused eosinophilic infiltration following viral re-exposure in mice [74, 75]. In contrast, at least in the case of SARS-CoV-1, immunization with RBD induced neutralizing antibodies in the absence of a TH2/TC2 response [76]. Vaccine adjuvants can stimulate TH1/TC1-favorable innate immunity [77, 82], as this is the case for multiple viral vectors, virus-like particles and mRNA containing nanoparticles. Finally, virus adaptation to the host has to be outcompeted. One might infer from our data that the currently protective immunodominant regions generating a TH1/TC1 profile may be the focus of the future antigenic drift of SARS-CoV-2.

Community-protective immunity can affect RNA virus evolution by selecting for new antigenic variants on the scale of years, as exemplified by the need for annual evaluation of influenza vaccines [78]. It is likely that pandemic SARS-CoV-2 evolution bears similarities with human influenza A virus evolution including accumulation of adaptive changes in the receptor binding domain (RBD), in which case vaccines would have to be updated regularly [78]. Therefore, to win the race against current virus strains and emerging variants, an expedited world-wide vaccination rollout ensuring an immunization *en masse* against relevant epitopes (and in particular the entire RBD region of the variants from Brazil, India, and South Africa) with vaccine formulations ensuring TH1/TC1 (rather than TH2/TC2) responses should outwit the COVID-19 pandemic. It will be of utmost interest to monitor the polarity and specificity elicited by current COVID-19 vaccines and to correlate these recognition profiles with failed immunization, reinfection and emergence of severe disease.

## SUPPLEMENTAL MATERIALS

### Material and methods

#### Patient and cohort characteristics

All clinical studies were conducted after written informed consent in accordance with Good Clinical Practice guidelines and the provisions of the Declaration of Helsinki. Cohorts’ characteristics are detailed in Table S1, Figure 1A and Figure S1. Two cohorts of cancer patients (from the pre-COVID-19 era and from the COVID-19 era) and three cohorts of healthy volunteers (from the pre-COVID-19 era and from the COVID-19 period including a cohort of vaccinees) were exploited to set up the translational research analyses. Peripheral blood mononuclear cells (PBMC) were provided by Gustave Roussy Cancer Campus (Villejuif, France) and IHU Méditerranée Infection (Marseille, France) (see Blood analyses section).

Contemporary clinical studies (COVID-19 era): 1/ ONCOVID clinical trial and regulatory approvals.

#### Principles

The protocol is available at https://clinicaltrials.gov/ct2/show/NCT04341207. Gustave Roussy Cancer Center sponsored the trial named ‘ONCOVID’ and collaborated with the academic authors on the trial design and on the collection, analysis, and interpretation of the data. Sanofi provided trial drugs. Protocol approval was obtained from an independent ethics committee (ethics protocol number EudraCT No: 2020-001250-21). ***Patients.*** ONCOVID eligible patients were all comer adults with advanced solid tumors or advanced hematological malignancies spontaneously presenting at Gustave Roussy between April 10^th^, 2020 and January 15^th^, 2021 (data cut-off for our analyses). ***Trial design.*** Cancer patients were screened for SARS-CoV-2 virus carriage by nasopharyngeal sampling at every hospital visit. The presence of SARS-CoV-2 RNA was detected by RT-qPCR assay in a BSL-2 laboratory. Asymptomatic and symptomatic patients (*i.e* presenting with fever (t°>38°C) and/or cough and/or shortness of breath and/or headache and/or fatigue and/or runny nose and/or sore throat, anosmia/ageusia) with a positive SARS-CoV-2 RT-qPCR test, shifted to the interventional phase (tailored experimental approach with Hydroxychloroquine and Azithromycin therapy in symptomatic SARS-CoV-2 positive subjects). Asymptomatic or symptomatic patients with negative SARS-CoV-2 RT-qPCR tests continued their standard of care anti-cancer treatments. Repeated RT-qPCR for SARS-CoV-2 on nasopharyngeal swabs and blood samples were performed to monitor the status for SARS-CoV-2 and the immune response, respectively, in COVID-19 positive and negative patients. The COVID-19 severity was defined based on oxygen, imaging and hospitalization criteria (WHO criteria). Unexposed and exposed individuals included in the ONCOVID trial during the first surge were followed-up by telephone interview at 12-months in order to record documented COVID-19 infections and degree of severity (WHO criteria) during the successive surges. ***Samples for translational research.*** PMBCs were isolated less than 8 hours after the blood collection (at patient inclusion and at every hospital visit) and kept frozen at-80°C. ***2/ PROTECT-Cov clinical trial and regulatory approvals*. *Principles.*** IHU Méditerranée Infection sponsored the trial named ‘PROTECT-Cov’ and collaborated with the academic authors on the trial design and on the collection, analysis, and interpretation of the data. Protocol approval was obtained from an independent ethics committee (ethics protocol number ANSM No: 2020-A01546-33). The trial was conducted in accordance with Good Clinical Practice guidelines and the provisions of the Declaration of Helsinki. All patients provided written informed consent. ***Subjects.*** PROTECT-Cov eligible subjects were members of the same family/home composed of two or more people and selected from the microbiology laboratory register on SARS-Cov-2 tests performed between March 23 and April 10, 2020. ***Trial design.*** Members of the same family/home who had at least one (a)symptomatic COVID-19 + case (RT-qPCR <35 Ct values for SARS-CoV-2 on nasopharyngeal swabs) and at least one member with negative RT-qPCR for SARS-CoV-2 (≥35 Ct) were screened. A telephone interview was conducted in order to confirm and complete the list of family circles in connection with the positive case. The compliant subjects finally selected were invited to come back to the IHU Méditerranée Infections hospital where they were included in the trial and had a blood test. ***3/ COVID-SER clinical trial and regulatory approvals*. *Principles.*** At the “Hospices Civils de Lyon”, France was conducted the trial named COVID-SER. Protocol approval was obtained from an independent ethics committee (the national review board for biomedical research, Comité de Protection des Personnes Sud Méditerranée, ID-RCB-2020-A00932-37). The clinical study was registered on ClinicalTrial.gov (NCT04341142). Written informed consent was obtained from all participants and the study. ***Subjects. COVID-SER*** eligible subjects were health care workers who received the Pfizer–BioNTech mRNA COVID-19 vaccine BNT162b2. All subjects with a positive SARS-CoV-2 RT-PCR prior vaccination and/or a positive serological result with Wantaï Ab total kit at the pre-vaccination visit were considered as convalescent. Blood sampling was performed before vaccination and 4 weeks after receiving 1 or 2 doses of vaccine for naive and convalescent health care workers respectively. According to French procedures, a written non-opposition to the use of donated blood for research purposes was obtained from healthy volunteers. The donors’ personal data were anonymized before transfer to our research laboratory. We obtained approval from the local ethical committee and the French ministry of research (DC-2008-64) for handling and conservation of these samples. Human biological samples and associated data were obtained from NeuroBioTec (CRB HCL, Lyon France, Biobank BB-0033-00046) and Virginie Pitiot.

**Clinical studies from the Pre-COVID-19 era: *1/ Series of patients with cancer:*** This cohort is composed of different IGR clinical trials. Patients were included and blood was collected and banked between 1999 and 2018 (Pre-COVID-19 era). ***Clinical studies:*** *1/* Patients with acute myeloid leukemia admitted in the Hematology Department of IGR in Villejuif, France, between March 2008 and March 2009 were included. The study was approved by the local ethics committee (Comité de Protection des Personnes (CPP) Hopital Bicêtre –CALEX protocol, n1 ID RCB 2007-A01074-49, date 29 February 2008). The main clinical and biological characteristics of the patients are summarized in [30] PMBC were isolated less than 8 hours after the blood collection (before chemotherapy) and were kept frozen at −80°C. *2/* Phase II vaccine trial immunizing cancer patients (diagnosed with inoperable Non-small cell lung cancer after induction with four cycles of platinum-based chemotherapy) with autologous DC-derived exosomes and cyclophosphamide (CTX) (Study code « Dex2 »: NCT01159288, date 19 December 2005) [28]. Eligible patients required at least stabilization of their disease prior to being on maintenance immunotherapy with sc injections of IFNγ-dendritic cell derived-exosomes loaded with MHC class I and class II-restricted cancer antigens. PBMCs were collected at baseline after the 4th cycle of platinum-based chemotherapy, before metronomic CTX followed by exosome injections and were kept frozen at −80°C. *3/* Phase I/II study of intradermal and subcutaneous immunization with the recombinant MAGE-3 protein in patients with MAGE-3 positive, measurable non visceral metastatic melanoma. The study was approved by the local ethics committee (Study code « LUD 99 003 »: N-CSET: 99/090/752, date 1 December 1999). Patients were required to have cutaneous melanoma with detectable cutaneous and/or lymph node metastasis, but no visceral metastasis (AJCC 1997 stage III or IVM1a). No chemotherapy, radiotherapy, or immunotherapy was allowed in the 4-week period before the first vaccination. PBMCs were collected at baseline before the first vaccination and were kept frozen at −80°C. 4/Adult patients with metastatic or locally advanced solid malignancy, measurable or evaluable disease who were refractory to standard therapy were eligible for the study (Phase I IMAIL-2 trial approved by the Kremlin Bicêtre Hospital Ethics Committee [no 07–019] and the Agence Française de Sécurité Sanitaire des Produits de Santé [no A70385–27; EudraCT N°:2007–001699–35 in 2007) [79]. They received Imatinib mesylate (IM) combined with increasing doses of IL-2. PMBCs were isolated less than 8 hours after the blood collection (at baseline before treatment) and were kept frozen at −80C. ***2/ Series of patients without cancer:*** Peripheral blood was obtained from healthy volunteers at the *Etablissement Français du Sang* (EFS, Paris France, n 18EFS031 date 24 September 2018).

#### Blood analyses

Blood samples (for serum and PBL) were drawn from patients enrolled in the different cohorts presented in the cohort description section above. Whole human peripheral blood was collected into sterile vacutainer tubes. ***Anti-SARS-CoV-2 immunoglobulins measurements*.** Serum was collected from whole blood after centrifugation at 600 g for 10 min at room temperature and transferred to −80°C freezer to await analysis. Serological analysis SARS-CoV-2 specific IgA, IgM and IgG antibodies were measured in 119 serum samples from 87 patients (Supplementary Material Figure 1) with The Maverick ™ SARS-CoV-2 Multi-Antigen Serology Panel (Genalyte Inc. USA) according to the manufacturer’s instructions. The Maverick™ SARS-CoV-2 Multi-Antigen Serology Panel (Genalyte Inc) is designed to detect antibodies to five SARS-CoV-2 antigens: nucleocapsid, Spike S1 RBD, Spike S1S2, Spike S2 and Spike S1 or seasonal HCoV-NL-63 nucleocapsid, -OC-43, −229E and -HK-U1 Spike in a multiplex format based on photonic ring resonance technology. This system detects and measures with good reproducibility changes in resonance when antibodies bind to their respective antigens in the chip. The instrument automates the assay. Briefly, 10µl of each serum samples were added in a sample well plate array containing required diluents and buffers. The plate and chip are loaded in the instrument. First the chip is equilibrated with the diluent buffer to get baseline resonance. Serum sample is then charged over the chip to bind specific antibodies to antigens present on the chip. Next, chip is washed to remove low affinity binders. Finally, specific antibodies of patients are detected with anti-IgG or -IgA or -IgM secondary antibodies. ***Isolation of peripheral blood mononuclear cells (PBMCs) from fresh blood sampling.*** Venous blood samples (10ml to 30ml) were collected in heparinized tubes (BD Vacutainer® LH 170 U.I., Dutscher, UK). On the same day, blood was processed in a biosafety level 2 laboratory at Gustave Roussy Institute, Villejuif, France, or in IHU Méditerranée Infection, Marseille, France. Peripheral blood mononuclear cells (PBMCs) were freshly isolated by LSM, Lymphocyte Separation Medium (Eurobio Scientific, France) density gradient centrifugation according to manufacturer’s instructions. (Leucosep tubes, Greiner; Biocoll, Bio&SELL). PBMCs were then collected, washed once with phosphate-buffered saline solution (PBS) and aliquoted in 1ml of cryopreservation medium (CryoStor®, STEMCELLS Technologies, USA) in cryovials (two cryovials per patient). Cryovials (Cryotube^TM^vials ThermoFisher Scientific, Denmark) were conserved for 24h at −80°C in a cryo-freezing container (Mr.Frosty^TM^,Thermo Fisher Scientific) before storage in liquid nitrogen.

#### Viral studies. Biosafety levels for in vitro experiments

Frozen PBMCs from patients with a confirmed negative RT-qPCR for SARS-CoV-2 genome at the time of blood drawing were processed in a biosafety level 2 laboratory at Gustave Roussy Institute, Villejuif, France. All samples from patients with positive RT-qPCR were processed in a biosafety level 3 laboratory at Henri Mondor Hospital, Créteil, France. When a patient was sampled at different timepoints, samples were processed together in the same laboratory. ***RT-qPCR analysis*.** SARS-CoV-2 diagnostic testing of clinical nasopharyngeal swabs or other samples by RT-qPCR was conducted from 14 March to 23 March 2020 at an outside facility using the Charité protocol. From the 23^th^ March 2020 testing was performed internally at the Gustave Roussy. The cycle thresholds were collected only for assays performed at Gustave Roussy. Nasopharyngeal swab samples were collected using flocked swabs (Sigma Virocult) and placed in viral transport media. SARS-CoV-2 RNA was detected using one of two available techniques at Gustave Roussy: the GeneFinder COVID-19 Plus Real*Amp* kit (ELITech Group) targeting three regions (*RdRp* gene, nucleocapsid and envelope genes) on the ELITe InGenius (ELITech Group) or the multiplex real-time RT-PCR diagnostic kit (the Applied Biosystems TaqPath COVID-19 CE-IVD RT-PCR Kit) targeting three regions (ORF1ab, nucleocapsid and spike genes) with the following modifications. Nucleic acids were extracted from specimens using automated Maxwell instruments following the manufacturer’s instructions (Maxwell RSC simplyRNA Blood Kit; AS1380; Promega). Real-time RT-PCR was performed on the QuantiStudio 5 Dx Real-Time PCR System (Thermo Fisher Scientific) in a final reaction volume of 20 μl, including 5 μl of extracted nucleic acids according to the manufacturer instruction. ***Viral lysates and their production.*** SARS-CoV-2 IHUMI2, IHUMI845, IHUMI846, IHUMI847 (early 2020 episode), IHUMI2096 (20A.EU2, B.1.160) and IHUMI2514 (20C, B.1.367) [80] IHUMI3076 (20I/501Y.V1, B.1.1.7), IHUMI3147 (20H/501Y.V2, B.1.351) and IHUMI3191 (20J/501Y.V3, P.1) strains were isolated from human nasopharyngeal swab as previously described [35] and grown in Vero E6 cells (ATCC CRL-1586) in Minimum Essential Medium culture medium (MEM) with 4% fetal calf serum (FCS) and 1% L-glutamine. Influenza strains H1N1 (0022641132) and H3N2 (8091056304) were isolated then produced from human nasopharyngeal swab in MDCK cells (ATCC CCL-34) in MEM with 10% FCS and 1% L-glutamine. All these clinical isolates were characterized by whole viral genome sequencing from culture supernatants. Coronavirus OC43 (ATCC vr-1558) was grown in HCT8 cells (ATCC CCL-244) in RPMI with 10% FCS. Coronavirus 229E (ATCC vr-740) was grown in MRC5 cells (ATCC CCL-171) in MEM with 10% FCS. All reagents for culture were from ThermoFisher Scientific and all cultures were incubated at 37°C under 5% CO_2_ without antibiotics. All viral strains were produced in 125 cm^2^ cell culture flasks. When destruction of cell monolayer reached approximately 80%, between 2 to 7 days according to cell line and viral strain, culture supernatant was harvested. After low speed centrifugation to remove cells and debris (700 x g for 10 min.) supernatants were filtered through 0.45 then 0.22 µm pore-sized filters. These viral suspensions were then inactivated for 1 hour at 65°C before use. Batches of scrapped control uninfected cells were rinsed twice in PBS, and then finally resuspended in 5 ml of PBS at 5.10^5^ cells/ml. All cells and antigens were tested negative for Mycoplasma before use.

#### In vitro stimulation assays

***Cross-presentation assay or peripheral blood lymphocyte stimulation with autologous monocyte derived-dendritic cells (DC).*** Frozen PBMCs were thawed, washed and resuspended in RPMI 1640 media (GIBCO). Viability and count were evaluated using a Vi-Cell XR Cell Counter (Beckman Coulter, Brea). PBMC were then cultured in RPMI 1640 supplemented with 10% human AB serum, 1mM Glutamine, 1% sodium pyruvate, 1% HEPES, 1% penicillin/streptomycin at a cell density of 0.5M cells/cm^2^ for 2 hours at 37°C, 5% CO_2_ and separated into adherent and non-adherent cell populations. Non-adherent cells, containing Peripheral Blood Lymphocytes (PBL), were collected and cultured 4 days at 37°C, 5% CO_2_ in IMDM medium (Sigma-Aldrich, UK), supplemented with 10% human AB serum (Institut de Biotechnologies Jacques Boy, France), 1mM Glutamine (GIBCO/ThermoFisher Scientific, UK) 1% Sodium Pyruvate (GIBCO/ThermoFisher Scientific, UK), 1% HEPES (GIBCO/ThermoFisher Scientific, UK), 1% penicillin/streptomycin (GIBCO/ThermoFisher Scientific, UK) and 200 UI/mL rhIL-2 (Miltenyi, Germany). The adherent cell population was cultured for 3 days, at 37°C, 5% CO_2_, in a mo-DC differentiating media containing RPMI 1640 supplemented with 10% human AB serum, 1mM Glutamine, 1% sodium pyruvate, 1% HEPES, 1% penicillin /streptomycin, 1000UI/mL rhGM-CSF (Miltenyi) and 250UI/mL human IFNα-2b (Introna, MSD France). At day 3, adherent cells were slowly detached by pipetting after 20 minutes of incubation at 4°C and 20.000 cells were seeded in 96 well round bottom plate and were pulsed, or not (control condition), overnight, at 37°C, 5% CO_2_, with 1/10 heat inactivated viral lysates, or their respective control (see viral lysates production section). Spinoculation (800g for 2h, Centrifuge 5810R, Eppendorf, Germany) was next performed to ensure synchronized capture of the viral particles by mo-DCs. For activation and maturation, adherent cells were stimulated with LPS (10 ng/mL, Thermofisher) and GM-CSF (1000UI/mL). After 6h, mo-DCs were washed twice to remove LPS from the media and 100 000 PBL/well were seeded onto mature mo-DCs. PBL alone served as negative control, and PBL stimulated with anti-CD3 and anti-CD28 microbeads (1μL/mL, Dynabeads T-Activator, InVitrogen) as a positive control. moDC-PBL co-culture was incubated at 37°C, 5% CO_2_ for 48h and supernatants were harvested and stored at −20°C.

#### Multiplex Cytokine Analysis or bead-based multiplex assays

moDC-PBL co-culture supernatants were analyzed using bead-based multiplex kit assays (MACSplex cytokine 12 human, Miltenyi) according to the manufacturer protocol. Briefly, 50uL of supernatant were used with a MACSPLEX Cytokine12 Capture Beads (Miltenyi, France) to measure the concentration of 12 cytokines (GM-CSF, IFN-α, IFN-γ, IL-10, IL-12, IL-17A, IL-2, IL-4, IL-5, IL-6, IL-9, TNF-a). Bead fluorescence was acquired on a CytoFLEX flow cytometer (Beckman Coulter) for samples processed at Gustave Roussy Institute and on a FACSAria Fusion (Becton Dickinson) for samples processed in the biosafety level 3 laboratory at Henri Mondor Hospital. FlowJo (Treestar, Ashland, OR, USA) software was used for analysis.

#### Peptide-based assay

Lyophilized peptides were dissolved in sterile water and used at 2µg/mL in RPMI 1640 glutamax media (GIBCO) supplemented with 1% penicillin/streptomycin (GIBCO). 185 single peptides were plated in duplicates in 96 well round bottom TPP treated culture plates. Peptide plates were then stored at −80°C until use. The day of the experiment, peptide plates were thawed at room temperature. Frozen PBMCs were thawed, washed and resuspended in RPMI 1640 media (GIBCO). Viability and count were evaluated using a Vi-Cell XR Cell Counter (Beckman Coulter, Brea). PBMCs were then plated in RPMI 1640 glutamax media (GIBCO) supplemented with 1% penicillin/streptomycin (GIBCO), with 200UI/mL rhIL-2 (Miltenyi) and 200UI/mL rhIL-15 (Miltenyi) at a cell density of 10×10^3^ cells and incubated with each peptide at 37°C, 5% CO_2_. PBMCs were stimulated with 60 ng/mL OKT-3 antibody (ThermoFisher Scientific, clone OKT3) or with 10µg/mL phytohemagglutinin as positive controls and PBMCs alone served as negative controls. After 6 hours, 20µL of human AB serum was added to each well and plates were incubated at 37°C, 5% CO_2_ for 6 additional days. On day 7, supernatants were harvested and frozen at −80°C. Concentration of IFNγ, IL-9, IL-5 and IL-17A in the culture supernatant was determined using a commercial ELISA kit (ELISA Max Deluxe set human IFN-γ, Biolegend).

#### Positivity threshold determination for cytokine concentration using multiplex assays and commercial ELISA assays

For multiplex assays (or ELISA), a 4 parameter logistic regression was fitted for each cytokine based on the APC mean fluorescent intensity(or Optical Density) of standard dilution samples using nlpr(v0.1-7). This model was then used to calculate the concentration of each sample of unknown concentration. For multiplex assays, a ratio was computed for each cytokine using the cytokine concentration measured in response to each virus (SARS-CoV-2, HCoV-229E, HCoV-OC43) divided by the median concentration of their respective biological controls (Vero 81, MRC5, HCT8). A positivity threshold was set up based on the ratio for each cytokine. A ratio of above 1.5 minimum was requested to consider the supernatant “positive” for a cytokine. When necessary, a higher threshold was set up as such, median cytokine concentration of the biological controls + 2 times the standard deviation of the biological control concentrations divided by the median concentration. For ELISA assays, a ratio was computed as the concentration of the sample divided by the mean concentration of the negative controls.

#### COVID IGRA Biomérieux assay utilized for the COVID-SER clinical trial vaccinees [42]

Fresh blood collected in heparanized tubes was stimulated for 22 hours at 37°C under 5% of CO2 with peptide pools targeting RBD (46 peptides) (bioMérieux,France) diluted in IFA solution (bioMérieux, France). The IFA solution was used as a negative control and a mitogen was used as a positive control. The peptides (15-mer) encompassed the whole RBD protein sequence and overlapped by 5-residues. The concentration of IFNγ in the supernatant was measured using the VIDAS automated platform (VIDAS® IFNγ RUO, bioMérieux). The positivity range was 0.08 −8 IU/mL and IFA positivity thresholds were defined at 0.08 IU/mL. The IFNγ response was defined as positive when the IFNγ concentration of the test was above threshold and the negative control was below threshold or when the IFNγ concentration of the test minus IFNγ concentration of the negative control was above threshold. All positive controls were ≥8 IU/mL.

Reagents: culture media, cytokines, ELISA and multiplex assays. *<u>PBMC isolation</u>*. Blood samples were collected in heparinized tubes BD Vacutainer® LH 170 U.I., from Dutscher (catalog reference: 367526), diluted in PBS 1X purchased from Eurobio Scientific (catalog reference: CS3PBS01-01) and transferred in Leucosep^TM-^ 50mL purchased from Greiner Bio-One (catalog reference: 227290). Blood was centrifuged using MF48-R Centrifuge from AWEL Industries (catalog reference: 20023001). PBMC were collected in Centrifuge tube 50mL TPP from Dutscher (catalog reference: 91050), washed with PBS 1X, resuspended in CryoStor^®^ CS10 purchased from STEMCELL^TM^ technologies (catalog reference: 5100-0001) and transferred in Cryotube^TM^ vials from ThermoFisher Scientific (catalog reference: 377267). Samples were finally conserved for 24h at −80°C in a cryo-freezing container Mr.Frosty^TM^ from Thermo Fisher Scientific before storage in liquid nitrogen. ***Cross-presentation assay or peripheral blood lymphocyte stimulation with autologous monocyte derived-dendritic cells (DC).*** Frozen PBMCs were thawed, washed and resuspended in RPMI Medium 1640 (1X) purchased from GIBCO (catalog reference: 31870-025). Counting and viability were evaluated using Vi-CELL^TM^ XR Cell Viability Analyzer from Beckman Coulter (catalog reference: AV13289).To separate adherent and non-adherent cell populations, PBMC were transferred in 6 or 24 well flat bottom Sterile tissue culture testplate TPP purchased from Dutscher (catalog reference: 92006 / 92024) and cultured in complex medium (Complex Medium 1) containing human AB serum (catalog reference: 201021334), purchased from Institut de Biotechnologies Jacques Boy France), RPMI Medium 1640 (1X) (catalog reference: 31870-025), Sodium Pyruvate (catalog reference: 11360-039), Penicillin /Streptomycin (catalog reference: 15140-122), L-Glutamine (200mM) (catalog reference: 25030-024) HEPES Buffer Solution (catalog reference: 15630-056), MEM NEAA (catalog reference: 1140-035), purchased from GIBCO/ThermoFisher Scientific. The Non-adherent fraction was cultured in another complex medium (Complex Medium 2) containing human AB serum, Iscove’s Modified Dulbecco’s Medium (catalog reference: I3390), from Sigma-Aldrich, Sodium Pyruvate (catalog reference: 11360-039), Penicillin/Streptomycin (catalog reference: 15140-122), L-Glutamine (200mM) (catalog reference: 25030-024) HEPES Buffer Solution (catalog reference: 15630-056), MEM NEAA (catalog reference: 1140-035) from GIBCO/ThermoFisher Scientific and Recombinant Human IL-2 (PHAR000306) from Gustave Roussy Institute pharmacy. The adherent fraction was differentiated into monocyte derived-dendritic cells (mo-DC) in a mo-DC differentiating media constituted with Complex Medium 1 supplemented with Recombinant Human GM-CSF Premium purchased from Miltenyi (catalog reference: 130-093-867) and human IFNα-2b (Introna) purchased from MSD (France) (catalog reference: PHAR008943). For activation and maturation, DCs were stimulated with LPS purchased from Invivogen (catalog reference:) and GM-CSF purchased from Miltenyi Biotec (catalog reference: 130-093-867). PBL and mo-DC were finally co-cultured into 96 well V bottom Sterile Nunc^TM^ plate, VWR purchased from Dutscher (catalog reference: 92097). For positive control, PBL were stimulated with Dynabeads^TM^ Human T-Activator CD3/CD28 purchased from GIBCO / ThermoFisher Scientific (catalog reference: 11131D). All cell cultures were performed at 37°C, 5% CO_2_ into Heraus^®^ incubator purchased from Kendro Laboratory Products, ThermoFisher Scientific (catalog reference: BB 6220) And supernatants were transferred into 96 well V bottom Sterile Nunc^TM^ plate, VWR purchased from Dutscher (catalog reference: 734-0491) and frozen. ***Peptide-based assay.*** 96 well V bottom Sterile Nunc^TM^ plate were coated with peptides at 2µg/mL in RPMI Medium 1640 (1X) (catalog reference: 31870-025) supplemented with 1% Penicillin/Streptomycin (catalog reference: 15140-122) and conserved at −80°C. PBMCs were then thawed and plated in plate containing peptides in RPMI Medium 1640 (1X) (catalog reference: 31870-025) supplemented with 1% Penicillin/Streptomycin (catalog reference: 15140-122) supplemented with Recombinant Human IL-15 Premium grade from Miltenyi biotec (catalog reference: 130-095-765) and Recombinant Human IL-2 (PHAR000306) from Gustave Roussy Hospital. For positive, PBMC were stimulated with functional grade CD3, OKT3 purchased from ThermoFisher Scientific (catalog reference: 16-0037-85). Cell cultures were then supplemented with human AB serum (catalog reference: 201021334) purchased from Institut de Biotechnologies Jacques Boy (France) and cultured at 37°C, 5% CO_2_. ***Cytokines monitoring.*** Supernatants from cultured cells from Cross-presentation assay were monitored using the MACSPlex Cytokine 12 Kit human purchased from Miltenyi Biotec (catalog reference: 130-099-169). Acquisitions and analyses were performed on CytoFLEX S purchased from Beckman Coulter (catalog reference: B75442)/FACSAria Fusion purchased from BDbiosciences and FlowJo Software from Treestar respectively. Whereas Supernatants from cultured cells from peptide-based assay were monitored using ELISA tests purchased from BioLegend: ELISA MAX^TM^ Deluxe Set Human IFN-γ (catalog reference: 430104) ELISA MAX^TM^ Deluxe Set Human IL-17 (catalog reference: 433914) and ELISA MAX^TM^ Deluxe Set Human IL-9 (catalog reference: 434705).

#### Rationale of peptide selection and peptide synthesis (Refers to **Table S6**)

Peptide selection and synthesis: the peptides from the spike and nucleocapsid proteins were selected by dividing the sequences of the SARS-CoV-2 spike protein (RefSeq ID QHD43416.1) and of the nucleocapsin protein (RefSeq ID QHD43423.2) in non-overlapping 15 amino acid segments. The peptides from the membrane protein were selecting by dividing the sequence of 2 potential immunogenic regions of the SARS-CoV-2 (RefSeq QHD43422.1) membrane protein in overlapping 15 amino acid segments. The peptides from the ORF8 and ORF10 proteins were selected by dividing the sequences of the SARS-CoV-2 ORF8 protein (RefSeq ID QHD43422.1) and of the ORF10 protein (RefSeq ID QHI42199.1) in overlapping 15 amino acid segments. The peptides from ORF3 and some for ORF8 were selected based on a previous study [81]. The SARS-CoV-1peptides were peptides found to be immunogenic in previous reported studies. The peptides were synthesized by peptides&elephants GmbH (Berlin, Germany). The peptide pools for the controls for Influenza, EBV and CMV were acquired from peptides&elephants GmbH (Berlin, Germany) order numbers LB01774, LB01361 and LB01232 respectively.

## Statistical analysis

All calculations, statistical tests, and data visualization were performed using R v4.0.3. All analyses were performed on independent samples, excepting when the presence of replicates is mentioned. The associations between continuous variables were evaluated using Spearman correlation. Group comparisons were performed using non-parametric test with the wilcox.test R function: the Wilcoxon-Mann-Whitney test for independent samples, and the Wilcoxon signed rank test for paired samples. The comparison of categorical data was performed using the Fisher exact test with the fisher.test R function. Hierarchical clustering was performed with the package hclust, using the euclidean distance. Linear and logistic regressions were performed with respectively the lm and the glm R base functions. A peptide set enrichment analysis was performed with the R package fgsea (version 1.14.0), using as statistic the t-value of the coefficient of univariable linear regressions of the logarithm-normalized IL-2 secretion on the different peptides. All hypothesis tests (including those of regression coefficients) were two-sided and considered as statistically significant when p<0.05. Graphical illustrations were drawn using the standard R packages dedicated to the data visualization (ggplot2, ggpubr, corrplot, complexheatmap, circlize, and Hmisc).

## Data Availability

The data that support the findings of this study are available on request from the corresponding author LZ.

## Acknowledgments

We thank the ET-EXTRA team (Biological Resource Center (NF 96-600) and the microbiology team for technical help. We thank the staff from health and safety of Gustave Roussy Cancer Campus for helping to set up the translational research studies. We are thankful to the Genalyte for their supportive help. We are thankful to Jeanne Magnan and Alexandre Trubert for their supportive help. Lyon COVID Study GROUP and authors thank the Hospices Civils de Lyon and by Fondation des Hospices Civils de Lyon and all the personnel of the occupational health and medicine department of Hospices Civils de Lyon who contributed to the samples collection.

## Contributions

L.D., L.Z., A.M. and F.B. conceived and designed the clinical trial. L.Z. conceived and designed the translational research, interpreted the data and formulated figure design. A.G., F-X.D., S.T., C.M., L.A., B.B., A.St., B.G., M.Mer., F.S., A.M., L.D. included patients in the clinical trial. J-E.F, I.L., A.C., M.Maz., C.T., Y.H., M.Pi., M.G., B.K., G.F., A.D., C.F., A.T. and J.M. carried out all the experiments. J-E.F., and D.D. performed all the statistical analyses. L.D., A.G. C.A-C-S. and G.M collected the clinical data. Sophie Assan provided the information from the HCL series with the contribution of BioMérieux. C.M, B.L.S., S.G. and S.C. analyzed and provided the clinical data from the IHU. E.C. and F.G. performed the RT-qPCR. A-G.G., C. A-C-S., M.Maz. and LD collected the biological data (ALC and Ct values). C.P. collected the samples. M.Maz., Y.H., E.P., C.F., G.F., C.T., A-G.G., B.K., A.T., and P.L. prepared the biological samples and provided some help for the experiments. B.L.S performed the viral cultures and provided viral lysates. A-A.B. helped with experiments performed in a BSL-3 confined environment. M.Mi. and G.G. performed the dosages of antibodies. E.d.S, MM and J.R.L designed the epitopes and prepared the peptides. All the authors advised for the interpretation of the data. L.Z. wrote the manuscript, with all authors contributing to writing and providing feedback.

## Funding

L.D. has received support by the Philanthropia Fondation Gustave Roussy. The Gustave Roussy sponsored clinical study on COVID-19 (ONCOVID; NCT NCT04341207 has been supported by the Fondation Gustave Roussy, the Dassault family, Malakoff Humanis, Agnès b., Izipizi, andRalph Lauren. A-G.G. and I.L. were supported by Fondation pour la Recherche Médicale (FRM). Lyon COVID study GROUP are Biomerieux employees or received grant from Bioméireux to perform experiments. L.Z. and G.K. were supported by RHU Torino Lumière (ANR-16-RHUS-0008), ONCOBIOME H2020 network, the SEERAVE Foundation, the Ligue contre le Cancer (équipe labelisée); Agence Nationale de la Recherche – Projets blancs; ANR under the frame of E-Rare-2, the ERA-Net for Research on Rare Diseases; Association pour la recherche sur le cancer (ARC); Cancéropôle Ile-de-France; FRM; a donation by Elior; the European Research Council (ERC); Fondation Carrefour; High-end Foreign Expert Program in China (GDW20171100085 and GDW20181100051), Institut National du Cancer (INCa); Inserm (HTE); Institut Universitaire de France; LeDucq Foundation; the LabEx Immuno-Oncology; the SIRIC Stratified Oncology Cell DNA Repair and Tumor Immune Elimination (SOCRATE); CARE network (directed by Prof. Mariette, Kremlin Bicêtre AP-HP), and the SIRIC Cancer Research and Personalized Medicine (CARPEM). G.I. and M.P. were supported by Italian Ministry of Health (grants Ricerca CorrenteLinea 1, 1 ‘Infezioni Emergenti e Riemergenti’, projects COVID-2020-12371675 and COVID-2020-12371817). M.Mi and G.G. were supported by ANR Flash COVID-19 program and ARS-CoV-2 Program of the Faculty of Medicine from Sorbonne University ICOViD programs (PI: G.G.). M.Mer. is a member of the Gates Foundation’s innate immunity advisory group and his work is supported by the Champalimaud Foundation through funds from grants UIDB/04443/2020, LISBOA-01-0145-FEDER-022231 and LISBOA-01-0246-FEDER-000007. Over the last 5 years: AM has been a Principal Investigator of Clinical Trials from the following companies: Roche/Genentech, BMS, Merck (MSD), Pfizer, Lytix pharma, Eisai, Astra Zeneca/Medimmune, Tesaro, Chugai, OSE immunotherapeutics, SOTIO, Molecular Partners, IMCheck, Pierre Fabre, Adlai Nortye. A.M. has been a Member of Clinical Trial Steering Committees for NCT02528357 (GSK), NCT03334617 (AZ) and a Member of Data Safety and Monitoring Board: NCT02423863 (Sponsor: Oncovir), NCT03818685 (Sponsor: Centre Léon Bérard). A.M. has been a compensated member of the following Scientific Advisory Boards: Merck Serono, eTheRNA, Lytix pharma, Kyowa Kirin Pharma, Novartis, BMS, Symphogen, Genmab, Amgen, Biothera, Nektar, Tesaro/GSK, Oncosec, Pfizer, Seattle Genetics, Astra Zeneca/Medimmune, Servier, Gritstone, Molecular Partners, Bayer, Partner Therapeutics, Sanofi, Pierre Fabre, RedX pharma, OSE Immunotherapeutics, Medicxi, HiFiBio, IMCheck, MSD, iTeos, Innate Pharma, Shattuck Labs, Medincell, Tessa Therapeutics, Deka Biosciences. A.M. has provided compensated Teaching/Speaker activities for Roche/Genentech, BMS, Merck (MSD), Merck Serono, Astra Zeneca/Medimmune, Amgen, Sanofi, Servier. AM has provided compensated Scientific & Medical Consulting for Roche, Pierre Fabre, Onxeo, EISAI, Bayer, Genticel, Rigontec, Daichii Sankyo, Imaxio, Sanofi/BioNTech, Molecular Partners, Pillar Partners, BPI, Faron, Applied Materials. AM has benefited of Non-Financial Support (travel expenses) from Astra Zeneca, BMS, Merck (MSD), Roche. A.M. is a shareholder of Pegascy SAS, Centessa Pharmaceuticals, HiFiBio, Shattuck Labs. A.M. has received pre-clinical and clinical research grants (Institutional Funding) from Merus, BMS, Boehringer Ingelheim, Transgene, Fondation MSD Avenir, Sanofi and Astra Zeneca. BLS has received founding from the French Government under the “Investments for the Future” programme managed by the National Agency for Research (ANR), Méditerranée-Infection 10-IAHU. E.D. reports grants and personal fees from ROCHE GENENTECH, grants from SERVIER, grants from ASTRAZENECA, grants and personal fees from MERCK SERONO, grants from BMS, grants from MSD, grant and personal fees from Boehringer and was supported by INCa 2018-1-PL BIO-06-1 outside the submitted work. J-P.S was supported by MSD Avenir and has received and personal fees from Roche, MSD, BMS, Lilly, AstraZeneca, Daiichi-Sankyo, Mylan, Novartis, Pfizer, PFO, LeoPharma and Gilead outside the submitted work.

### The authors declare the following competing interests

L.Z. and G.K. are cofounders of everImmune, a biotech company devoted to the use of commensal microbes for the treatment of cancers. A.G., A.B.T. and A.M. as part of the Drug Development Department (DITEP) are Principal/sub-Investigator of Clinical Trials for Abbvie, Adaptimmune, Aduro Biotech, Agios Pharmaceuticals, Amgen, Argen-X Bvba, Arno Therapeutics, Astex Pharmaceuticals, Astra Zeneca, Astra Zeneca Ab, Aveo, Bayer Healthcare Ag, Bbb Technologies Bv, Beigene, Bioalliance Pharma, Biontech Ag, Blueprint Medicines, Boehringer Ingelheim, Boston Pharmaceuticals, Bristol Myers Squibb, Bristol-Myers Squibb International Corporation, Ca, Celgene Corporation, Cephalon, Chugai Pharmaceutical Co., Clovis Oncology, Cullinan-Apollo, Daiichi Sankyo, Debiopharm S.A., Eisai, Eisai Limited, Eli Lilly, Exelixis, Forma Tharapeutics, Gamamabs, Genentech, Gilead Sciences, Glaxosmithkline, Glenmark Pharmaceuticals, H3 Biomedicine, Hoffmann La Roche Ag, Incyte Corporation, Innate Pharma, Institut De Recherche Pierre Fabre, Iris Servier, Janssen Cilag, Janssen Research Foundation, Kura Oncology, Kyowa Kirin Pharm. Dev., Lilly France, Loxo Oncology, Lytix Biopharma As, Medimmune, Menarini Ricerche, Merck Kgaa, Merck Sharp & Dohme Chibret, Merrimack Pharmaceuticals, Merus, Millennium Pharmaceuticals, Molecular Partners Ag, Nanobiotix, Nektar Therapeutics, Nerviano Medical Sciences, Novartis Pharma, Octimet Oncology Nv, Oncoethix, Oncomed, Oncopeptides, Onyx Therapeutics, Orion Pharma, Oryzon Genomics, Ose Pharma, Pfizer, Pharma Mar, Philogen S.P.A., Pierre Fabre Medicament, Plexxikon, Rigontec Gmbh, Roche, Sanofi Aventis, Sierra Oncology, Sotio A.S, Syros Pharmaceuticals, Taiho Pharma, Tesaro, Tioma Therapeutics, Wyeth Pharmaceuticals France, Xencor, Y’s Therapeutics, Research Grants from Astrazeneca, BMS, Boehringer Ingelheim, Janssen Cilag, Merck, Novartis, Pfizer, Roche, Sanofi. Non-financial support (drug supplied) from Astrazeneca, Bayer, BMS, Boringher Ingelheim, Johnson & Johnson, Lilly, Medimmune, Merck, NH TherAGuiX, Pfizer, Roche. N.L. reports to be a Speaker at Jazz Pharmaceutical. Lyon COVID study GROUP are Biomerieux employees or received grant from Bioméireux to perform experiments. F.S. reports consulting fees from Pfizer, BMS, MSD, Roche, Pierre Fabre Oncology, Leo Pharma, Bayer, Mylan/Viatris, Mundi Pharma, Vifor Pharma, Biogaran, Helsinn. D.D. reports consulting fees from Chugai and Roche. E.D. reports grants and personal fees from ROCHE GENENTECH, grants from SERVIER, grants from ASTRAZENECA, grants and personal fees from MERCK SERONO, grants from BMS, grants from MSD, grant and personal fees from Boehringer outside the submitted work. F.B. reports personal fees from Astra-Zeneca, Bayer, Bristol-Myers Squibb, Boehringer– Ingelheim, Eli Lilly Oncology, Hoffmann–La Roche Ltd, Novartis, Merck, MSD, Pierre Fabre, Pfizer and Takeda, outside the submitted work. J-C.S. was a full-time employee of AstraZeneca between September 2017 and December 2019. J-C.S. reports consultancy: Relay Therapeutics, Gritstone Oncology and shares: Gritstone, AstraZeneca, Daiichi-Sankyo, outside the submitted work. G.G. is a member of the scientific board of Luxia Scientific and reports consultancy for Pileje and Luxia Scientific, outside the submitted work.

## Supplementary materials

**Figure S1.**
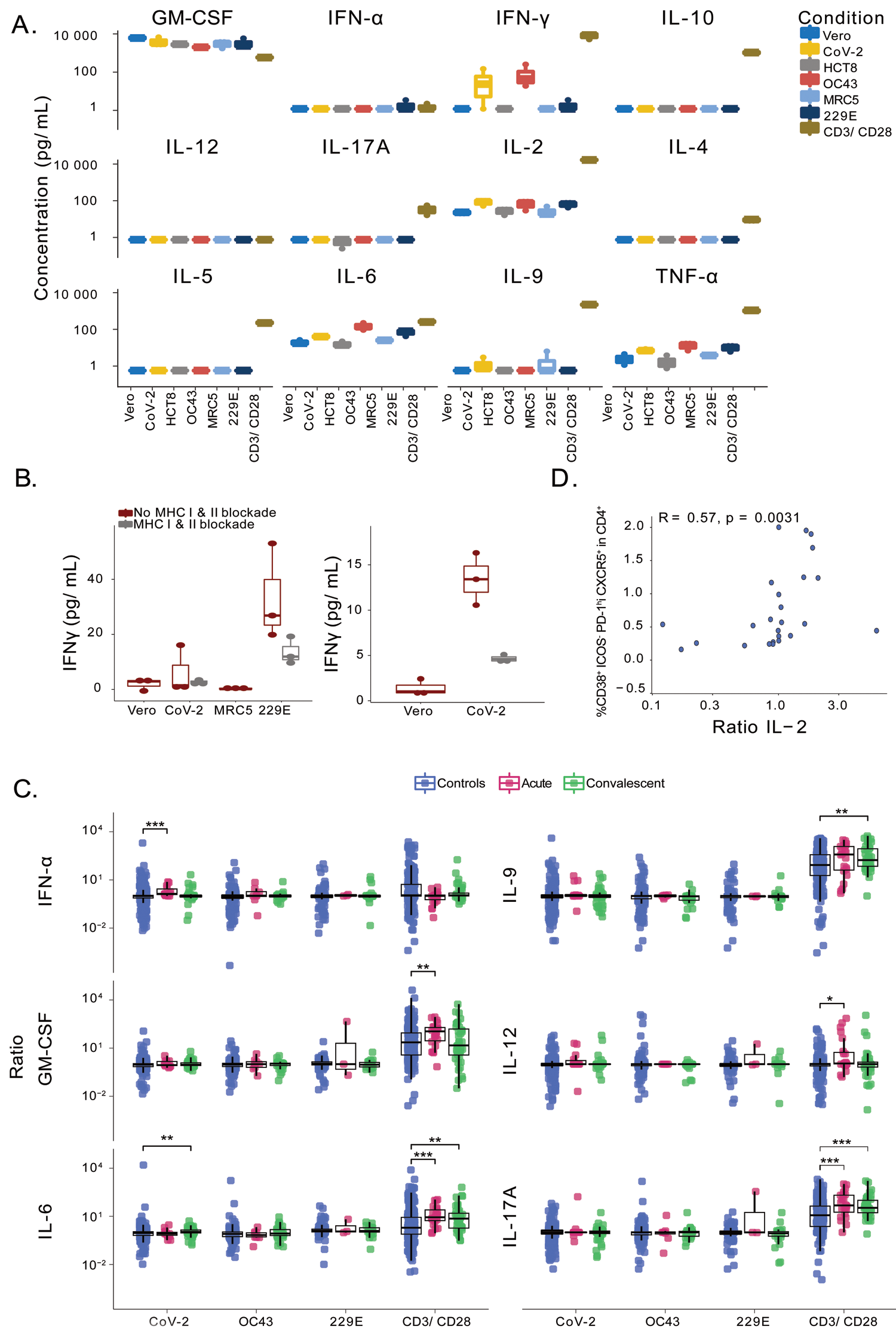
Cross-presentation *in vitro* assay at the acute and convalescent phase of COVID-19. **A.** Prototypical example of the result of the assay detailed in Figure 1B and M&M for one COVID-19 negative cancer patient. Each box plot represents the absolute values of cytokine secretion of two replicate wells in the 12 plex flow cytometric assay, in the 48hr supernatants of PBL stimulated with DC pulsed with SARS-CoV-2 versus Vero E6 (negative control) and the other common cold coronaviruses and their respective control cell line (negative controls). Each dot represents each replicate well for each stimulatory condition for one patient. Positive control: PBL stimulated with anti-CD3 and anti-CD28 antibody coated beads. **B.** MHC class I and class II neutralization experiments. Neutralizing anti-HLA-ABC and HLA-DR, DP, DQ antibodies (W6/32 & Tu39) were used to block the specific PBL reactivities in the above assay. **C.** Idem as in Figure 1D showing the other 6 cytokine ratios. **D**. Spearman correlation between SARS-CoV-2-specific IL-2 release and non-activated TFH. Group comparisons were performed using two-sided Wilcoxon-Mann-Whitney test and the asterisks indicate statistically significant differences (\**p*<0.05, \*\**p*<0.01, \*\*\**p*<0.001, \*\*\*\**p*<0.0001).

**Figure S2.**
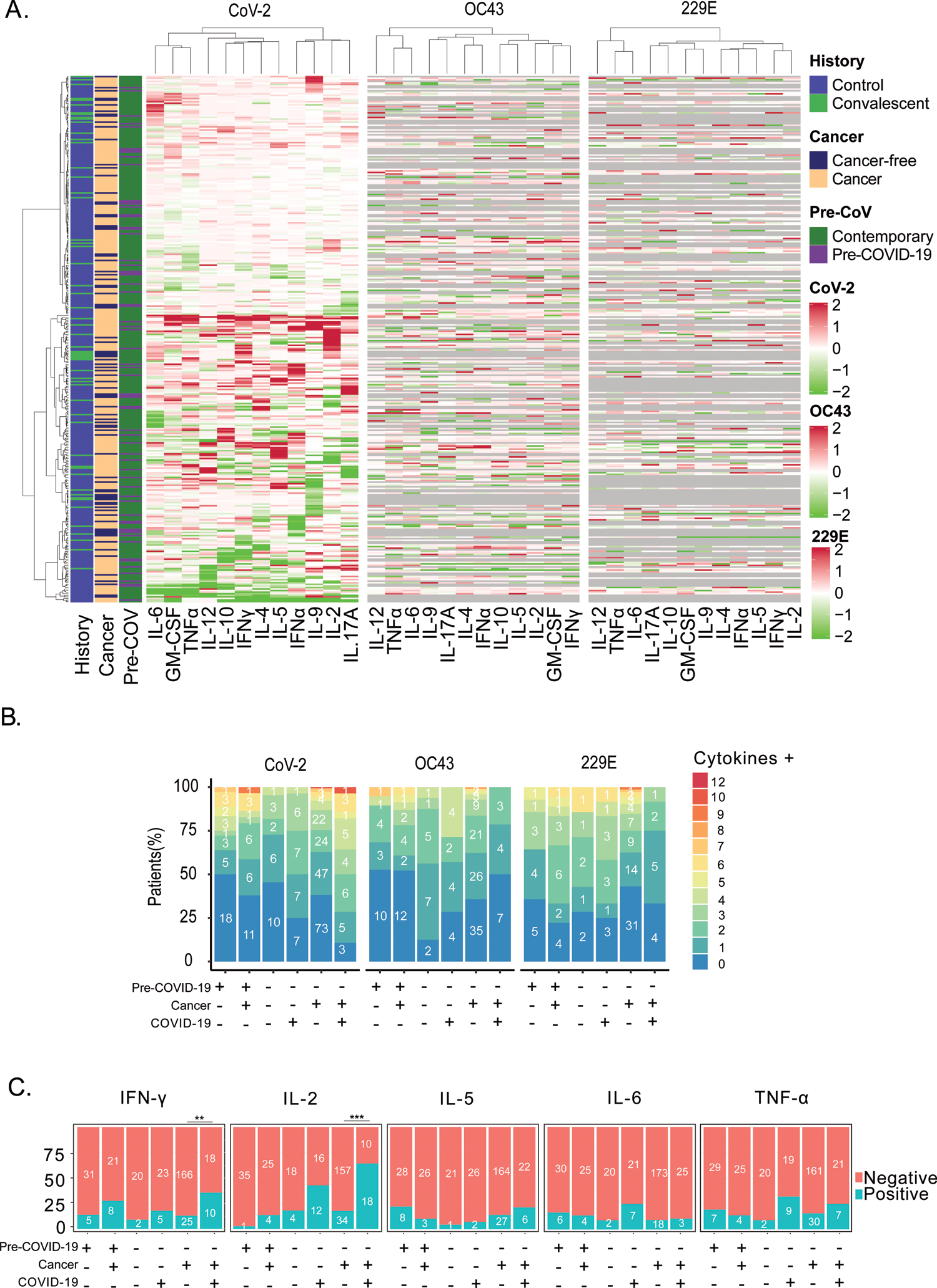
Detailed SARS-CoV-2 and CCC-specific cytokine release for convalescent COVID-19 patients compared with unexposed individuals. A. Non supervised hierarchical clustering of SARS-CoV-2 and CCC-specific cytokine release in all patients. Heatmap of cytokine release for SARS-CoV-2 or CCC in the cross-presentation assay performed at the convalescent phase of COVID-19 (n=54) and in unexposed individuals (n=304), aligning all 12 cytokines and all subject categories. **B.** Polyfunctional T cell responses among subjects from various cohorts. Individuals who exhibited SARS-CoV-2-specific release of one or several cytokines are enumerated in each bar. Each bar indicates a category of patients or subjects as described in Figure 1A. **C.** Percentage and number of patients in each cohort (Pre-COVID19 era (yes (+)/no(-)), Cancer (yes (+)/no(-) and COVID-19 (yes (+)/no(-)) who had a SARS-CoV-2-specific cytokine release (for the 5 statistically significant cytokines at the convalescent phase) compared with VeroE6 (Control, n=304; Convalescent, n=54). Asterisks indicate statistically significant differences of SARS-CoV-2-specific cytokine release proportions between two groups determined using Fisher exact test (\**p*<0.05).

**Figure S3.**
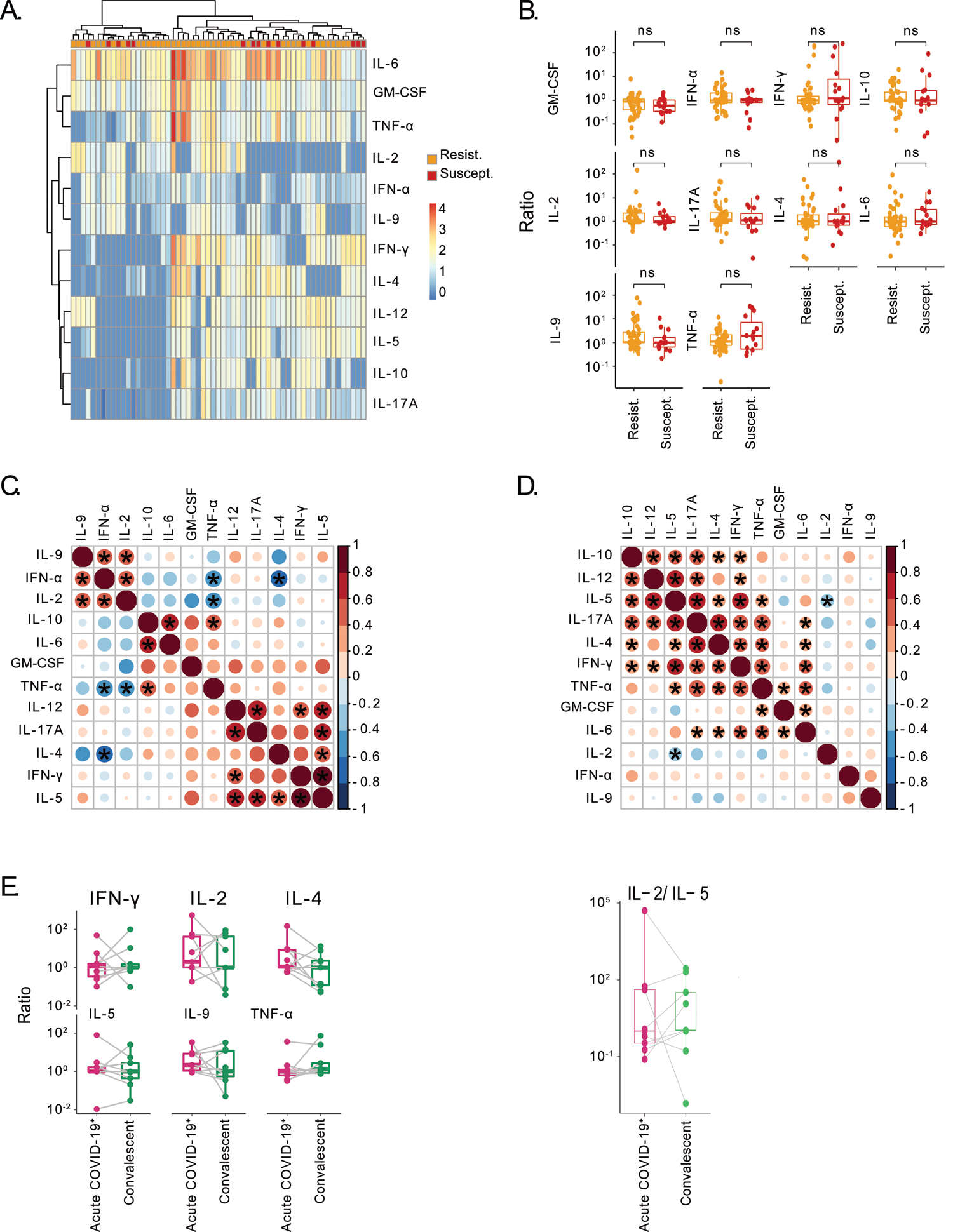
TH1/TC1 differentiation patterns in susceptible *versus* resistant individuals. A-B. Unsupervised hierarchical clustering of SARS-CoV-2-specific cytokine release. Heatmap of cytokine release in the cross-presentation assay performed during the first surge of the pandemic in unexposed individuals (n=60), aligning cytokines in the two subject categories, susceptible (persons who got infected during the second or the third surge of the pandemic) *versus* resistant (contact) individuals. Group comparisons were performed using a two-sided Wilcoxon-Mann-Whitney test. **C-D.** Spearman correlation matrices between cytokine levels in susceptible (n=19, C) *versus* resistant (n=41, D) individuals, respectively. The asterisks indicate statistically significant differences (*p*<0.05). **E**. Dynamic study of the stability of the TH1/TH2 profile in individuals that were followed up at two time points. Ratio of cytokine release at the acute and convalescent phase (left panel) and corresponding IL-2/IL-5 ratio (right panel) in 5 cancer patients. Two-sided Wilcoxon-Mann-Whitney test did not reveal significant difference between both time points.

**Figure S4.**
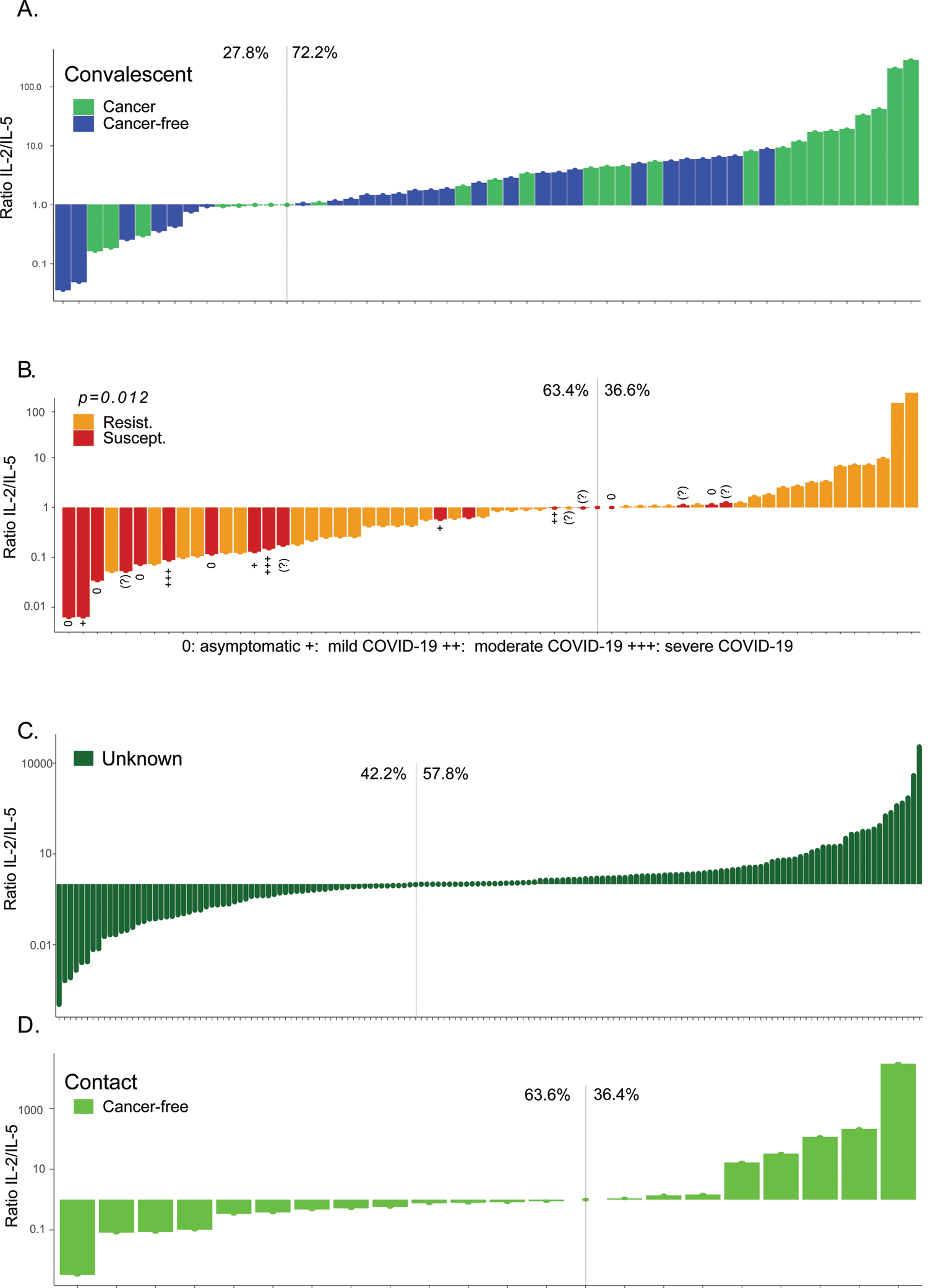
Waterfall plots indicating the IL-2/IL-5 ratio in all patient or individual groups. Waterfall plot between IL-2 and IL-5 ratio of cytokine release in the Figure 1B IVS assay in all patients during the first surge of the pandemic depicting cancer (green) versus cancer free (blue) COVID-19+ convalescent patients (A), resistant (orange) versus susceptible (red) (B), locked down (or unknown) subjects (dark green, n=301) and healthy individuals in contact with their COVID-19+ family members (light green) (D). Each bar represents one patient. Proportion of patients exhibiting an IL-2/IL-5 ratio superior or inferior to 1 is indicated in each panel. Clinical conditions are annotated as 0, +, ++, +++ for asymptomatic, mild, moderate, and severe COVID-19 severity, respectively. Refer to Figure 2F where percentages are compared inbetween groups.

**Figure S5.**
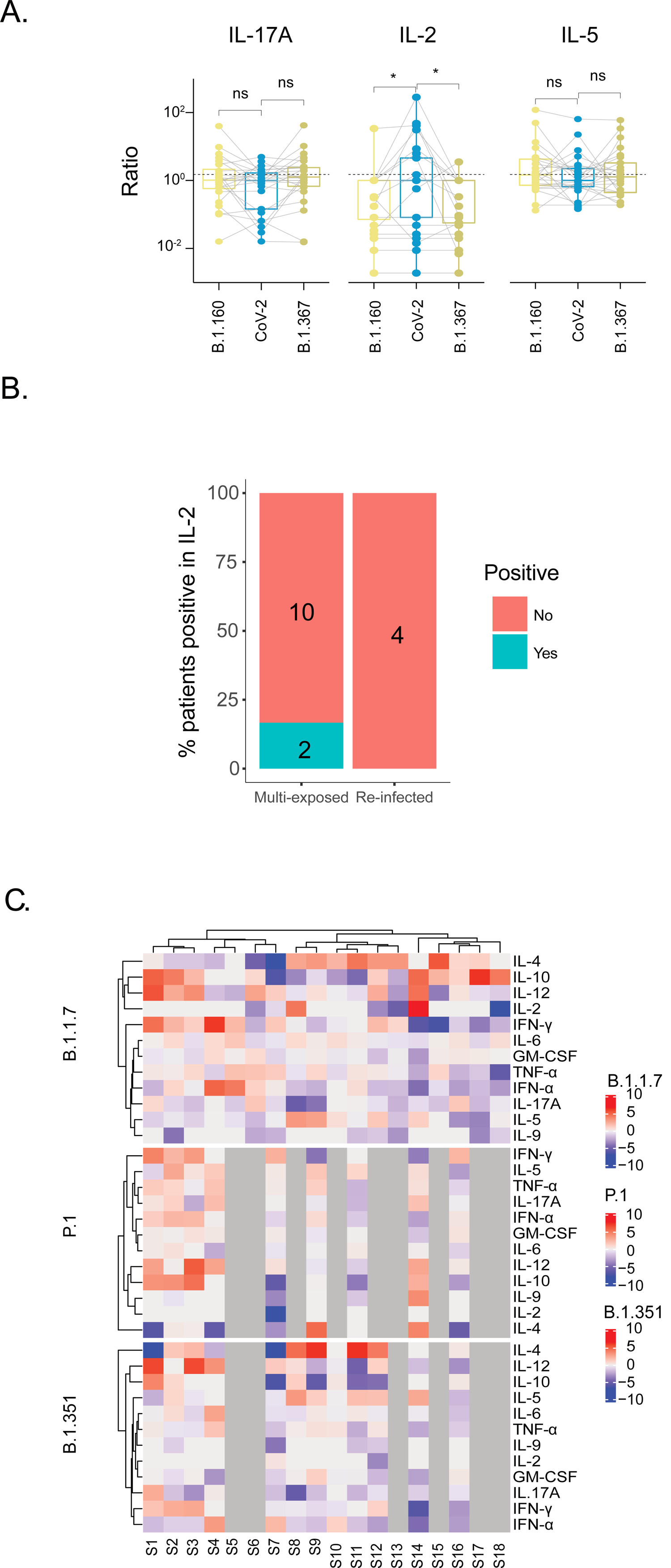
Cross-presentation assays using viral variants. A. Cytokine ratio in the cross-presentation assay detailed in Figure 1B, using the original strain IHUMI846 (early 2020 episode) (CoV-2 in (A)), versus the Danish mink (B.1.160, 20A.EU2, GH) and North African (B1.367, 20C, GH) strains in 25 control individuals. Statistical comparisons were performed using paired two-sided Wilcoxon-Mann-Whitney test and the asterisks indicate statistically significant differences (\**p*<0.05, \*\**p*<0.01, \*\*\**p*<0.001, \*\*\*\**p*<0.0001). **B.** Percentage and number of patients in Fig2J right panel who had a SARS-CoV-2-specific IL-2 release. Of note, only 4 re-infected patients could be tested because DC could not be differentiated into monocytes in the others to allow the cross-presentation assay. **C**. Similar experimental setting as in Figure 1B but loading lysates from various SARS-CoV-2 strains (original one IHUMI846 (early 2020 episode), as well as IHUMI3076 (UK variant, B.1.1.7), IHUMI3147 (South African variant, B.1.351) and IHUMI3191 (Brazilian variant, P.1) strains isolated from human nasopharyngeal swab as previously described [35]. Heatmap of the intensity of the ratios between means of cytokine levels (of experimental replicates) obtained from PBL in coculture with DC+ IHUMI846 *versus* DC+ each variant.

**Figure S6.**
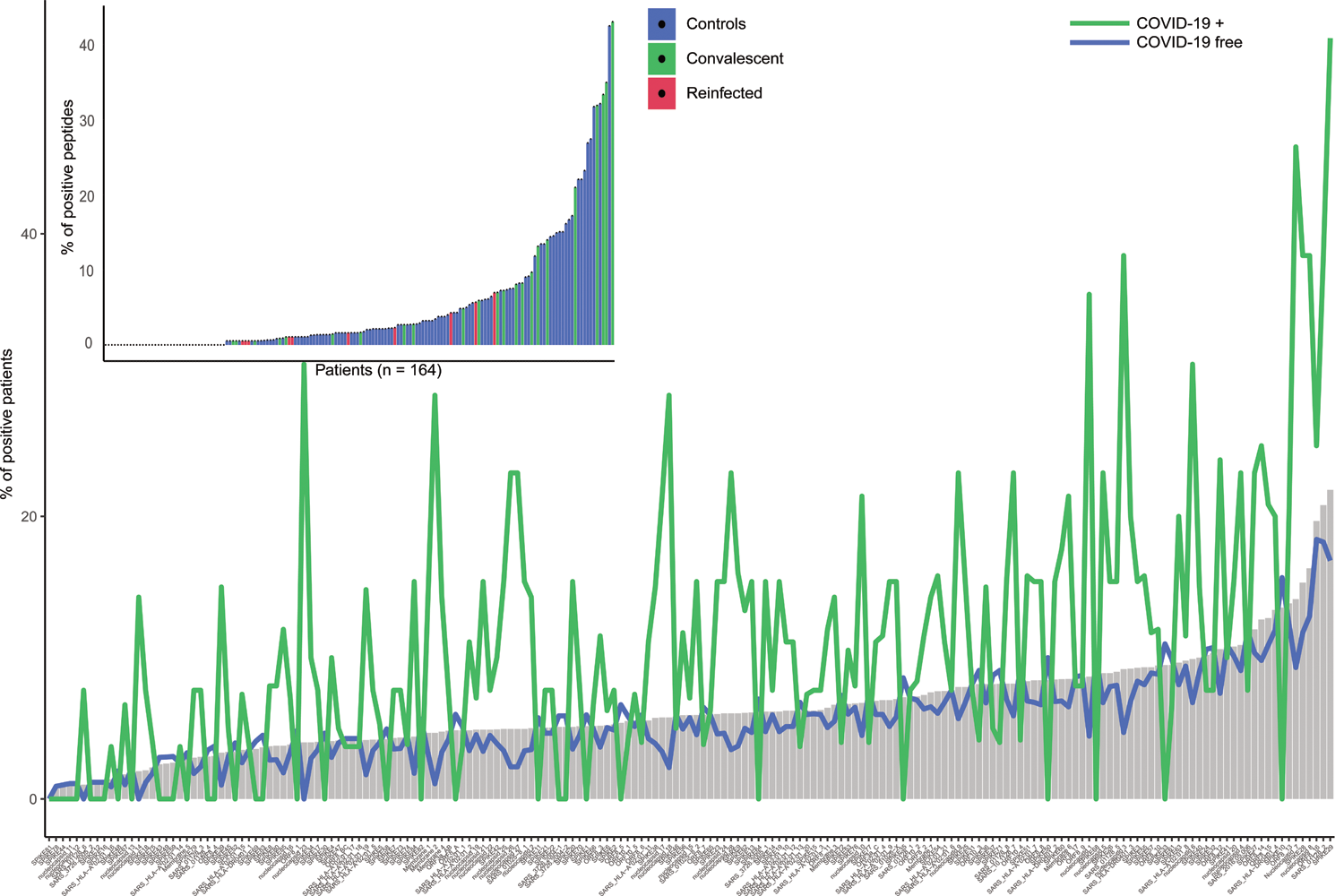
Percentages of patients reacting to each peptide and of peptides recognized by each individual in various groups or cohorts. Bar plot of the percentage of patients recognizing a given peptide. Blue and green lines represent the frequency of peptide recognition in control versus convalescent individuals respectively. Inset. Bar plot of the percentage of positive peptides in the IFNγ ELISA per patient. The color code indicates patient category (blue for control, green for convalescent COVID-19, purple for re-infected patients).

**Figure S7.**
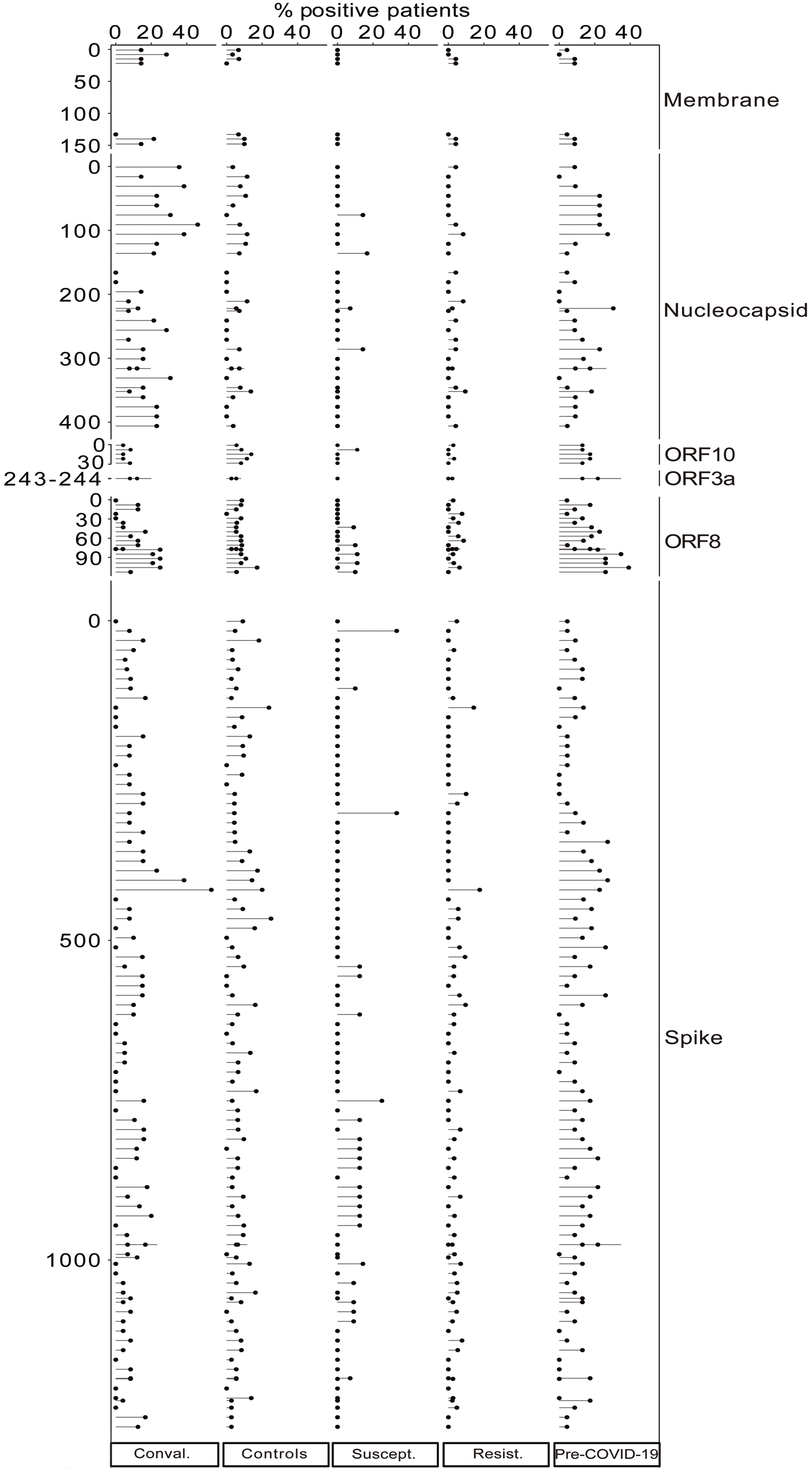
Peptide recognition mapping in various categories of individuals. Mean frequencies of IFNγ ELISA positivity in the peptide IVS assay (described in Figure 3A) for each indicated protein of the ORFeome in each subject category, for each individual peptide (each line corresponding to one peptide) in 159 subjects.

**Figure S8.**
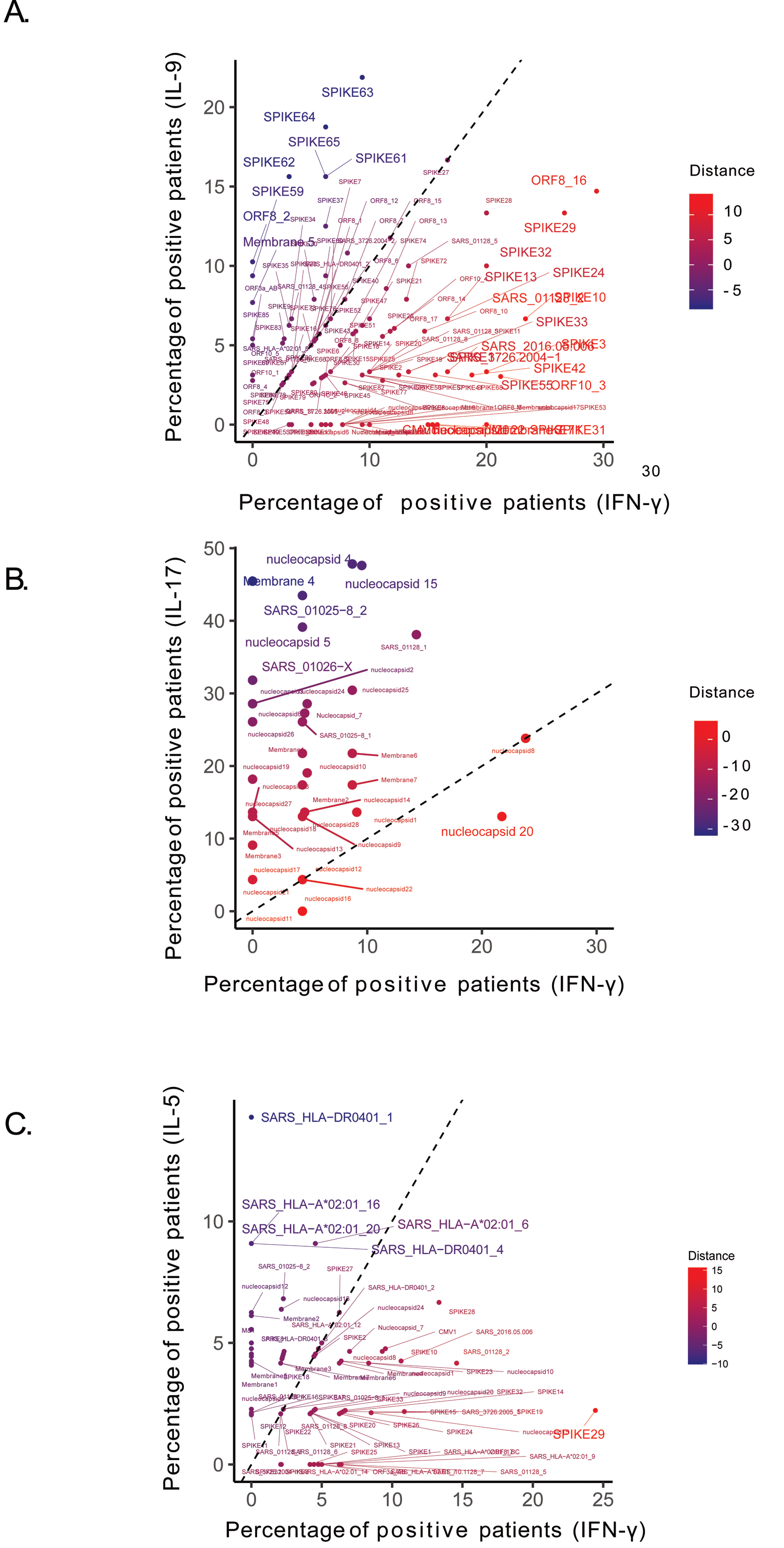
Correlation of secretory profiles for each peptide in the IVS assay. Scatter plot of peptides according to the percentage of patients reacting in IFNγ or IL-9 secretion (n=43, A) IL-17 (n=23, B) or IL-5 (n=64, C). The distance to the diagonal is indicated by a gradient of color.

**Figure S9.**
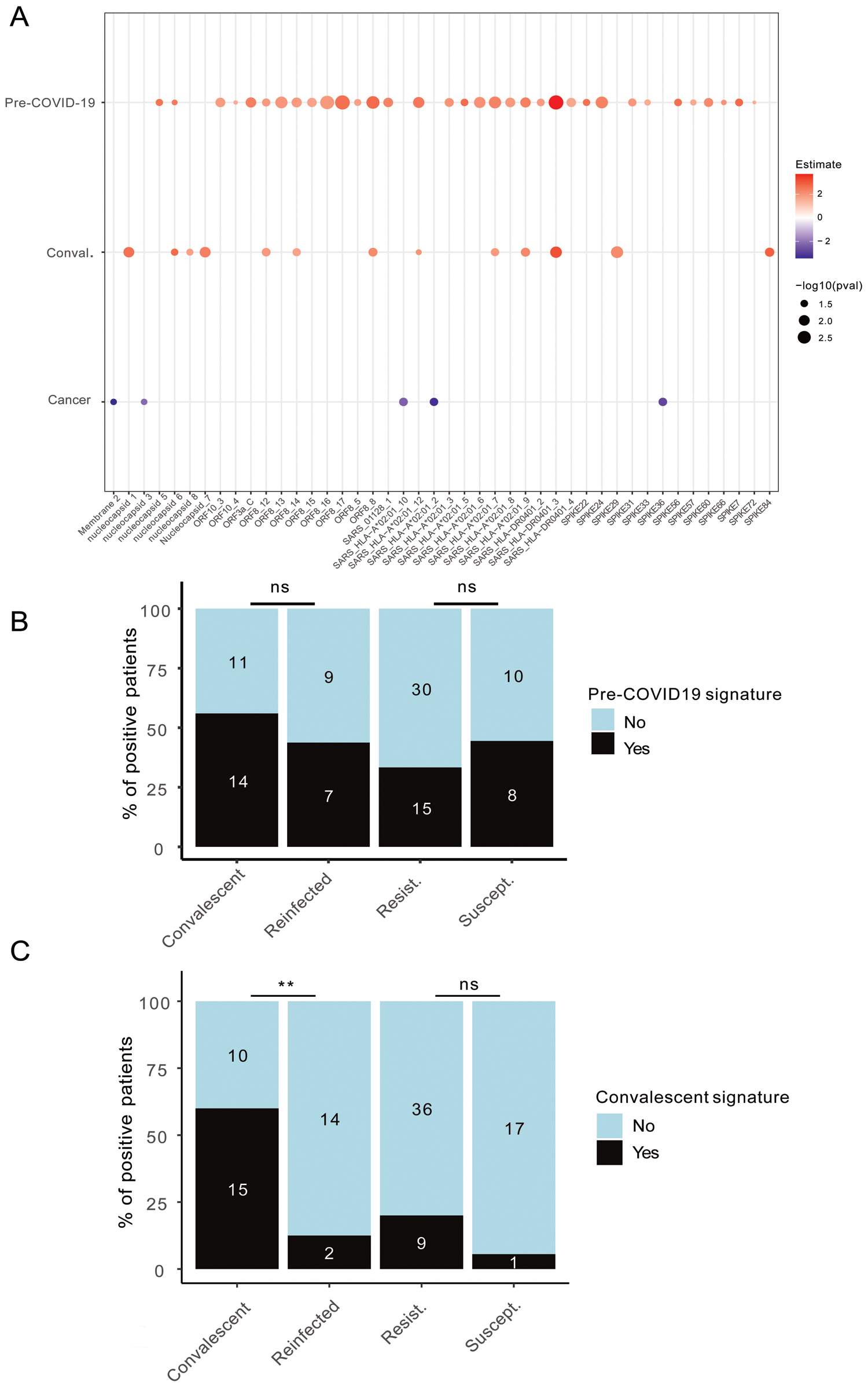
Logistic regression analyses identifying cohort-specific fingerprints of T cell repertoires. A. Statistically significant peptide signatures in the peptide-based IVS assay (Figure 3A) using a multivariable logistic regression analysis adjusted for period (pre-COVID-19 era or contemporary patients), COVID-19 history and cancer (refer to Table S2). The left column shows variables, and the x axis indicates the significant peptides (pval<0.05). The magnitude of the log (Odd Ratio) is indicated in the red/blue color code while that of the p-value is represented by the circle size. **B-C.** Percentages of patients recognizing at least one peptide from the pre-COVID-19 (B) or convalescent (C) signature identified in the logistic regression analyses of panel A in the IFNγ ELISA in the peptide IVS assay. Fisher’s exact test to compare the number of patients positive for each signature between groups (\**p*<0.05, \*\**p*<0.01).

## Supplemental Tables

**Table S1.**
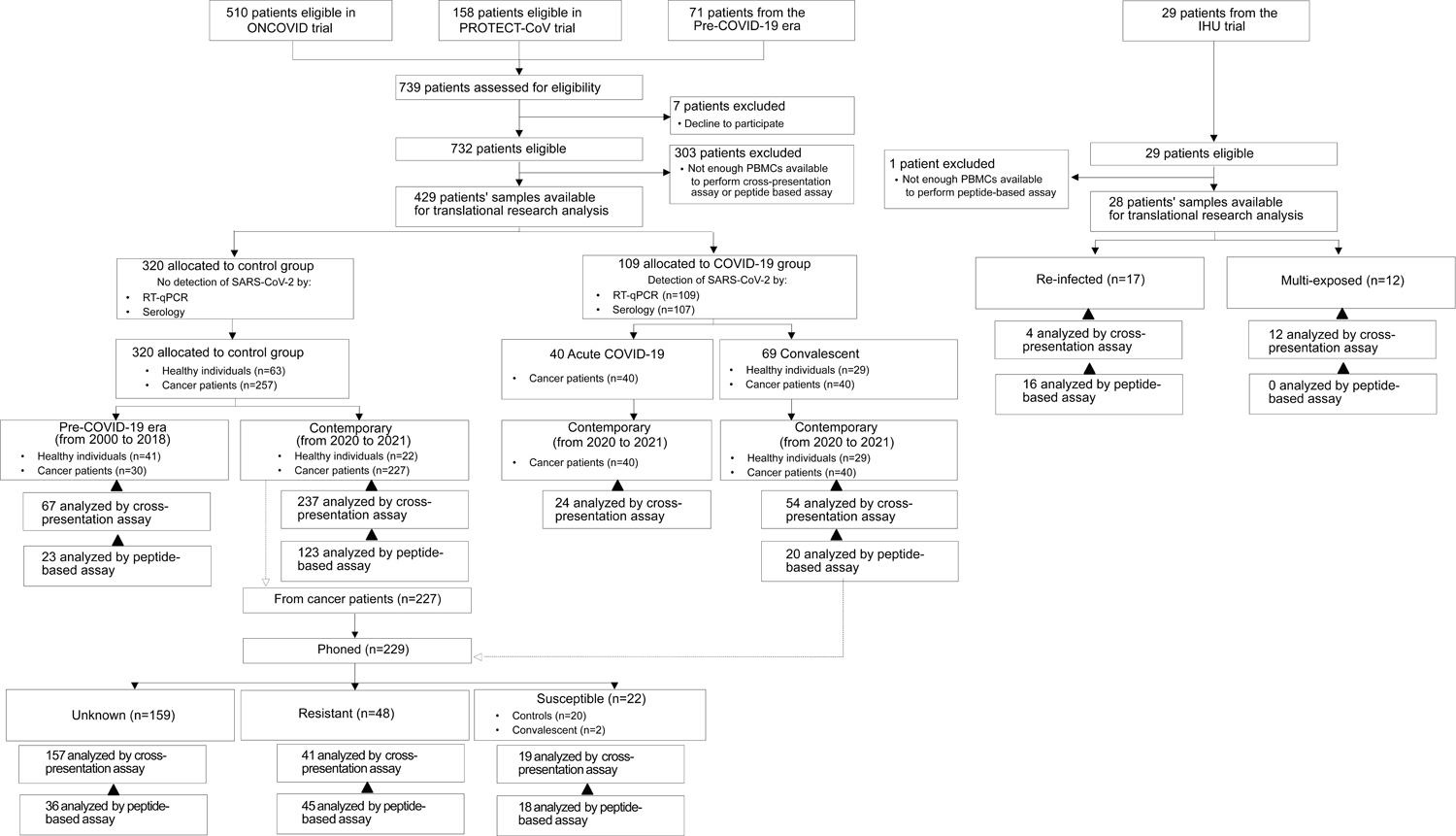
**Consort diagram.** Flow diagram showing the enrollment of subjects, their allocation to different cohorts and how they are analyzed in the trial.

**Table S2.**
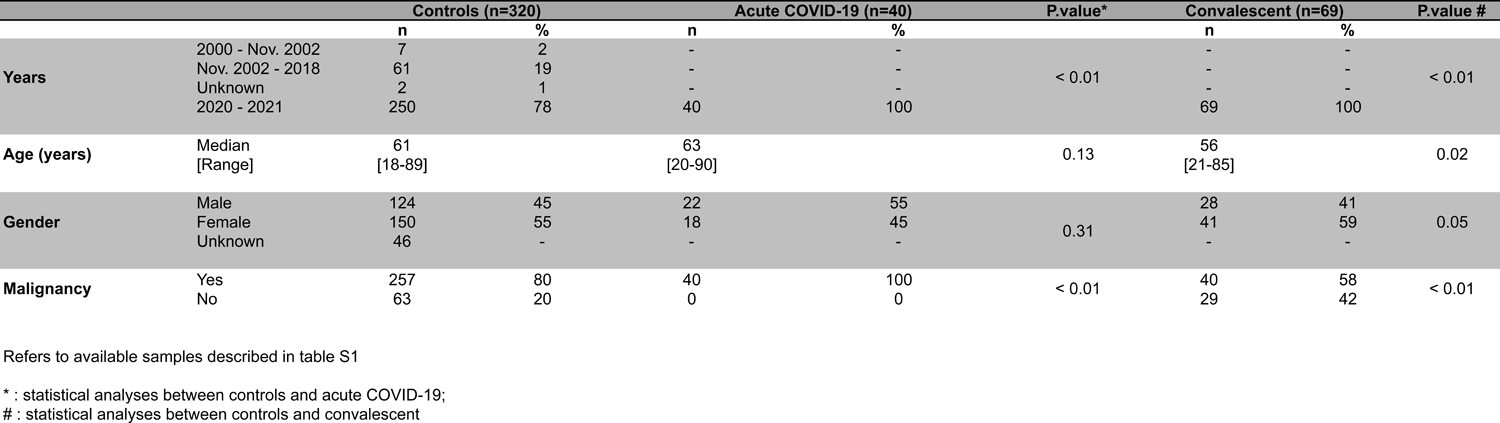
**Characteristics of cancer patients and healthy volunteers.**

**Table S3.** **Characteristics of family members during the 2020 lock down.**

|  |  | Total |  | Contact (n=22) |  | Convalescent (n=29) |  | P.value |
| --- | --- | --- | --- | --- | --- | --- | --- | --- |
| Age (years) | Median<br>[Range] | 46<br>[17-75] |  | 39<br>[17-75] |  | 49<br>[20-62] |  | 0.69 |
| Gender |  | n | % | n | % | n | % | 0.40 |
|  | Male | 26 | 51 | 13 | 59 | 13 | 45 |  |
|  | Female | 25 | 49 | 9 | 41 | 16 | 55 |  |
| Number of contacts per case within the family |  |  |  |  |  |  |  |  |
| 1-3 |  |  |  | 5 | 23 | 1 | 3 | < 0.01 |
| 4-6 |  |  |  | 14 | 64 | 15 | 52 |  |
| 7-9 |  |  |  | 1 | 5 | 8 | 28 |  |
| 10-12 |  |  |  | 2 | 9 | 0 | 0 |  |
| > 12 |  |  |  | 0 | 0 | 5 | 17 |  |
| Number of SARS-CoV-2 positive individuals in the family |  |  |  |  |  |  |  |  |
| 0 |  |  |  | 4 | 18 | 0 | 0 | < 0.01 |
| 1-3 |  |  |  | 18 | 82 | 18 | 62 |  |
| 4-6 |  |  |  | 0 | 0 | 5 | 17 |  |
| 7-15 |  |  |  | 0 | 0 | 6 | 21 |  |
| Household size |  |  |  |  |  |  |  |  |
| 0 |  |  |  | 2 | 9 | 0 | 0 | < 0.01 |
| 1-2 |  |  |  | 0 | 0 | 5 | 17 |  |
| 3-5 |  |  |  | 18 | 82 | 16 | 55 |  |
| 6-8 |  |  |  | 2 | 9 | 8 | 28 |  |
| Household living space (m²) |  |  |  |  |  |  |  |  |
| < 70 |  |  |  | 7 | 32 | 6 | 21 | 0.16 |
| 70-100 |  |  |  | 3 | 14 | 11 | 38 |  |
| > 100 |  |  |  | 12 | 55 | 12 | 41 |  |
| Number of rooms in the household |  |  |  |  |  |  |  |  |
| 2-3 |  |  |  | 10 | 45 | 7 | 24 | 0.22 |
| 4-5 |  |  |  | 6 | 27 | 14 | 48 |  |
| 6-8 |  |  |  | 6 | 27 | 8 | 28 |  |
| Number of individuals using personal protective equipment |  |  |  | 13 | 59 | 18 | 62 | 1 |
| Number of individuals respecting barriers gestures |  |  |  | 16 | 73 | 22 | 76 | 1 |

**Table S4.**
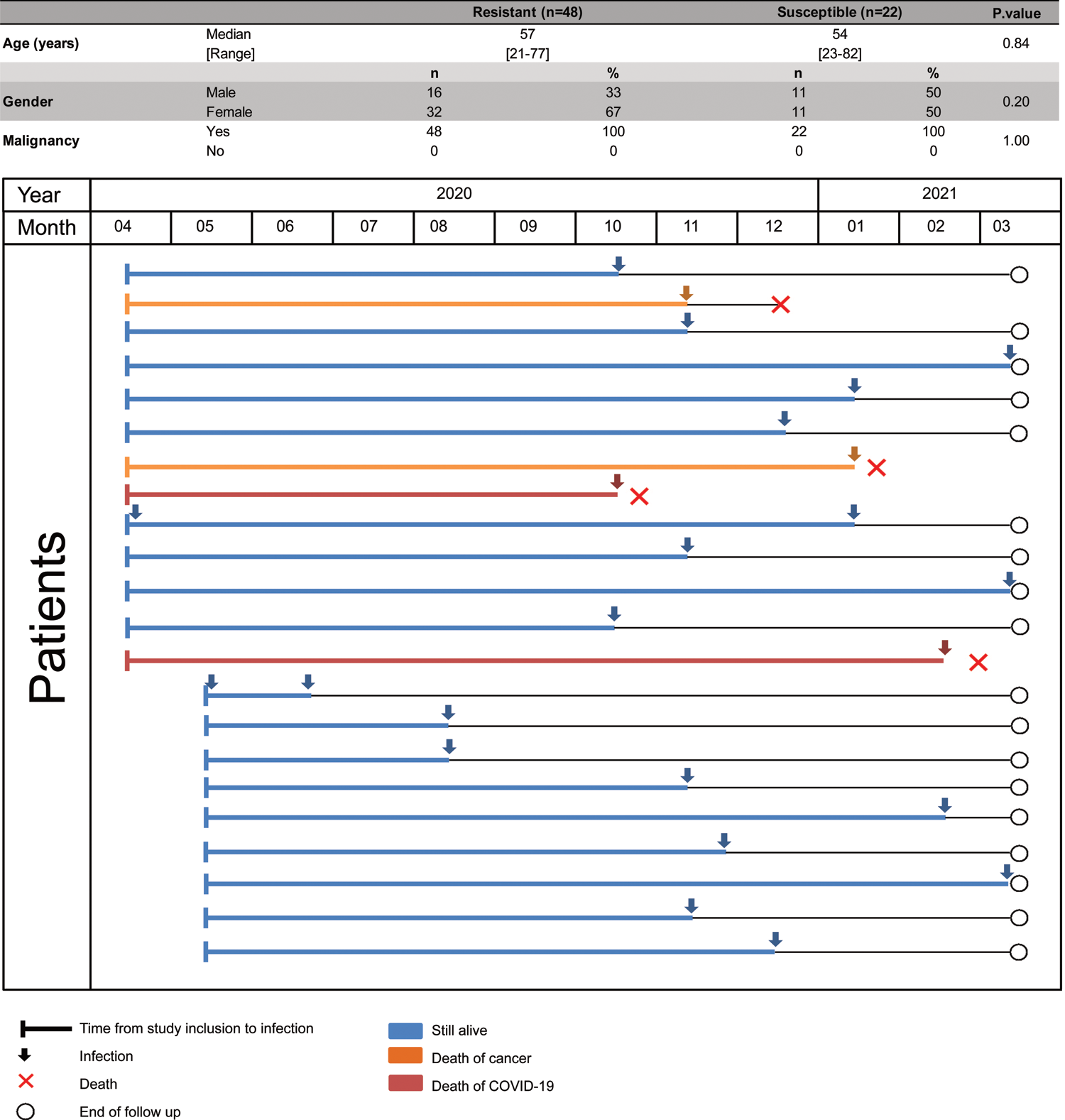
Characteristics of contact (resistant) and infected (susceptible) cancer patients and corresponding swimmer plot for the outcome of the susceptible cases. Individual swimmer plots for each susceptible individual (*i.e*. who got infected during the second or the third surge of the pandemic) depicting patients still alive (blue), patients dead from cancer, and patients dead from COVID-19 (red) after infection indicated with an arrow.

|  |  | Resistant (n=48) |  | Susceptible (n=22) |  | P.value |
| --- | --- | --- | --- | --- | --- | --- |
| Age (years) | Median | 57 |  | 54 |  | 0.84 |
|  | [Range] | [21-77] |  | [23-82] |  |  |
|  |  | n | % | n | % |  |
| Gender | Male | 16 | 33 | 11 | 50 | 0.20 |
|  | Female | 32 | 67 | 11 | 50 |  |
| Malignancy | Yes | 48 | 100 | 22 | 100 | 1.00 |
|  | No | 0 | 0 | 0 | 0 |  |

Time from study inclusion to infection
Infection
Death
End of follow up
Still alive
Death of cancer
Death of COVID-19

**Table S5.** **Characteristics of multi-exposed cases and re-infected cancer-free individuals.**

|  |  | Re-infected (n=17) |  | Multi-exposed cases (n=12) |  | P.value |
| --- | --- | --- | --- | --- | --- | --- |
| Age (years) | Median | 37 |  | 48 |  | 0.08 |
|  | [Range] | [16-78] |  | [28-69] |  |  |
|  |  | n | % | n | % |  |
| Gender | Male | 7 | 41 | 6 | 50 | 0.72 |
|  | Female | 10 | 59 | 6 | 50 |  |
| Comorbidities | Yes | 5 | 31 | - | - | - |
|  | No | 11 | 69 | - | - |  |
|  | Unknown | 1 | - | - | - |  |
| Type of comorbidities | Asthma | 3 | 60 | - | - | - |
|  | Obesity | 2 | 40 | - | - |  |
|  | Type 2 diabetes | 2 | 40 | - | - |  |
|  | Cancer |  | 20 | - | - |  |
|  | HIV* | 1 | 20 | - | - |  |
|  | HBP** | 2 | 40 | - | - |  |
| Time to reinfection (mo.) | < 03 | 2 | 17 | - | - | - |
|  | 03 - 06 | 7 | 58 | - | - |  |
|  | > 06 | 3 | 25 | - | - |  |
|  | Unknown | 5 | - | - | - |  |
| Second surge variants | B.1.177 | 1 | - | - | - | - |
|  | B.1.160 | 6 | - | - | - |  |
|  | B.1.351 | 1 | - | - | - |  |
|  | B.1.1.7 | 2 | - | - | - |  |
|  | Unknown | 7 | - | - | - |  |

Refers to eligible patients described in table S1
\*Human Immunodeficiency Virus
\*\*High Blood Pressure

**Table S6.**
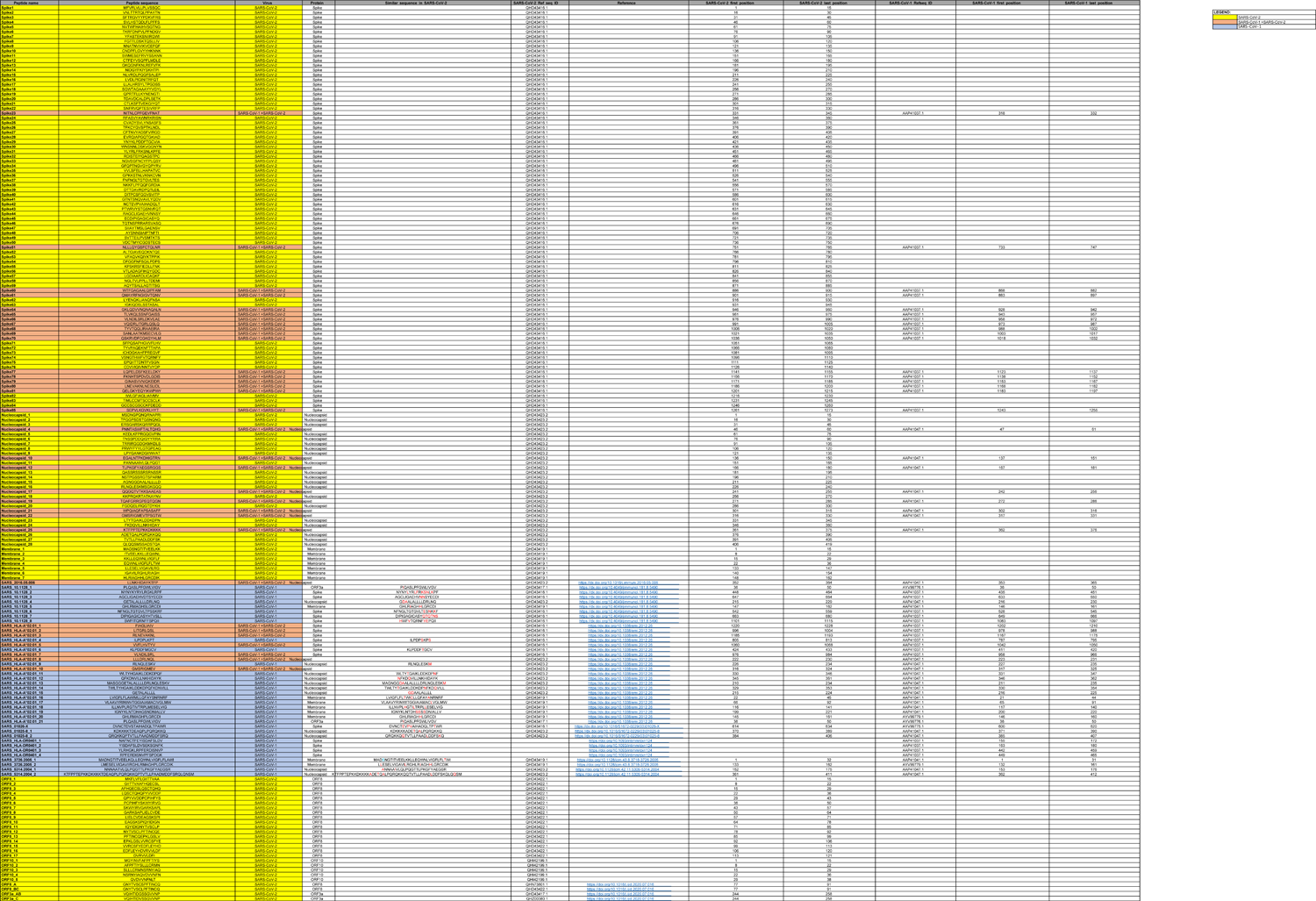
**SARS-CoV-1 and CoV-2 orfeome peptide list: sequences and positions.**

**Table S7.**
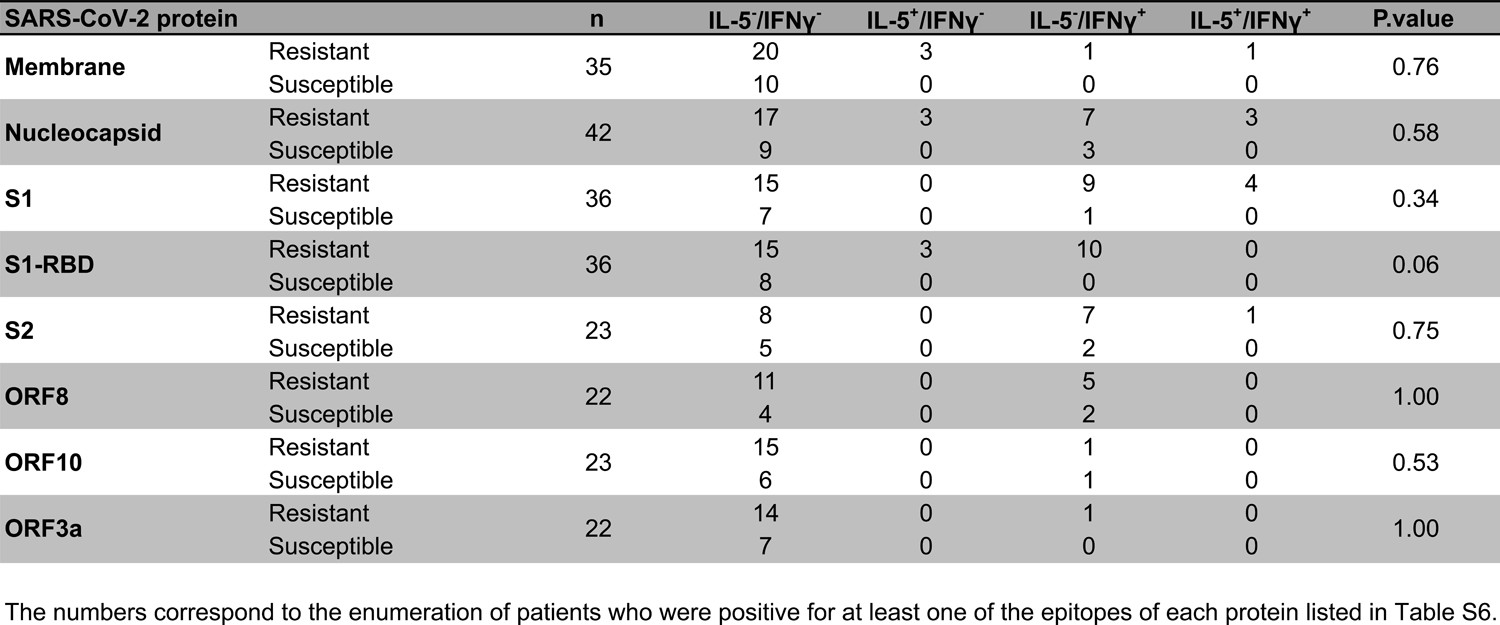
Peptide-specific T cell polarization determined by IL-5 or IFNγ quantification.

| SARS-CoV-2 protein | | n | IL-5<sup>-</sup>/IFNγ<sup>-</sup> | IL-5<sup>+</sup>/IFNγ<sup>-</sup> | IL-5<sup>-</sup>/IFNγ<sup>+</sup> | IL-5<sup>+</sup>/IFNγ<sup>+</sup> | P.value |
| --- | --- | --- | --- | --- | --- | --- | --- |
| Membrane | Resistant | 35 | 20 | 3 | 1 | 1 | 0.76 |
|  | Susceptible |  | 10 | 0 | 0 | 0 |  |
| Nucleocapsid | Resistant | 42 | 17 | 3 | 7 | 3 | 0.58 |
|  | Susceptible |  | 9 | 0 | 3 | 0 |  |
| S1 | Resistant | 36 | 15 | 0 | 9 | 4 | 0.34 |
|  | Susceptible |  | 7 | 0 | 1 | 0 |  |
| S1-RBD | Resistant | 36 | 15 | 3 | 10 | 0 | 0.06 |
|  | Susceptible |  | 8 | 0 | 0 | 0 |  |
| S2 | Resistant | 23 | 8 | 0 | 7 | 1 | 0.75 |
|  | Susceptible |  | 5 | 0 | 2 | 0 |  |
| ORF8 | Resistant | 22 | 11 | 0 | 5 | 0 | 1.00 |
|  | Susceptible |  | 4 | 0 | 2 | 0 |  |
| ORF10 | Resistant | 23 | 15 | 0 | 1 | 0 | 0.53 |
|  | Susceptible |  | 6 | 0 | 1 | 0 |  |
| ORF3a | Resistant | 22 | 14 | 0 | 1 | 0 | 1.00 |
|  | Susceptible |  | 7 | 0 | 0 | 0 |  |

The numbers correspond to the enumeration of patients who were positive for at least one of the epitopes of each protein listed in Table S6.

**Table S8.**
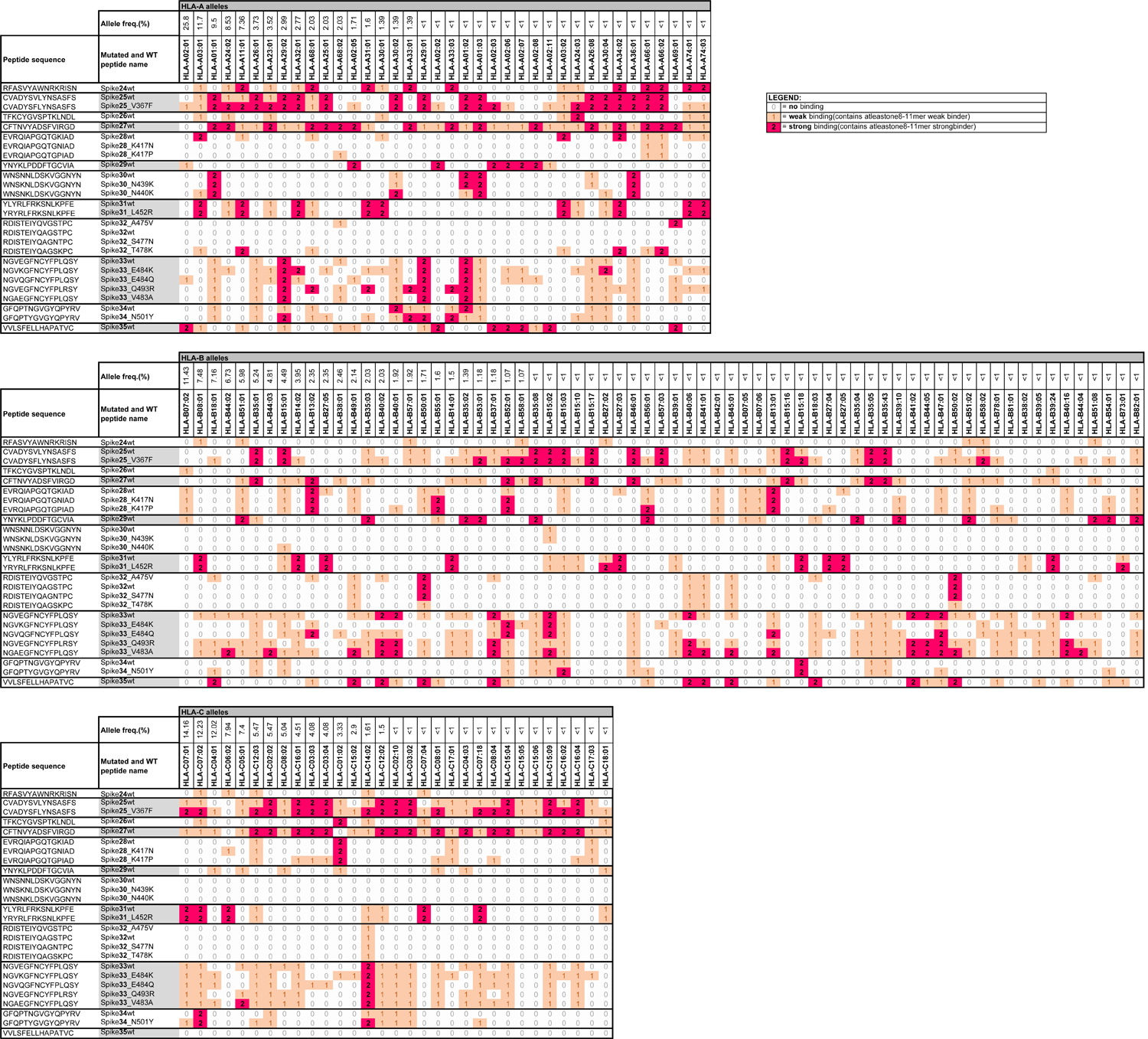
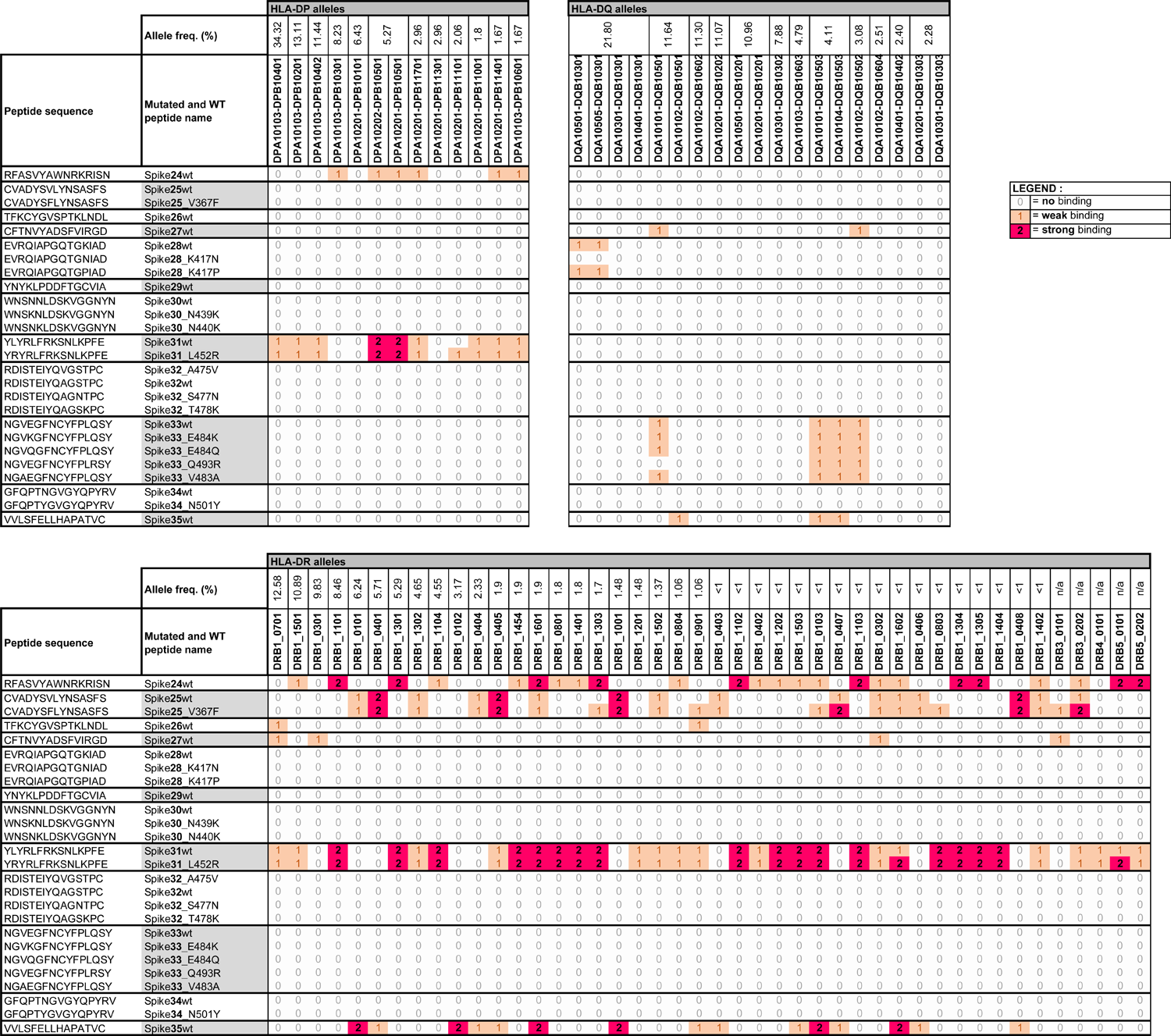
HLA binding affinities of the S1-RBD peptides using the NetMHCpan algorithm

**Table S9.**
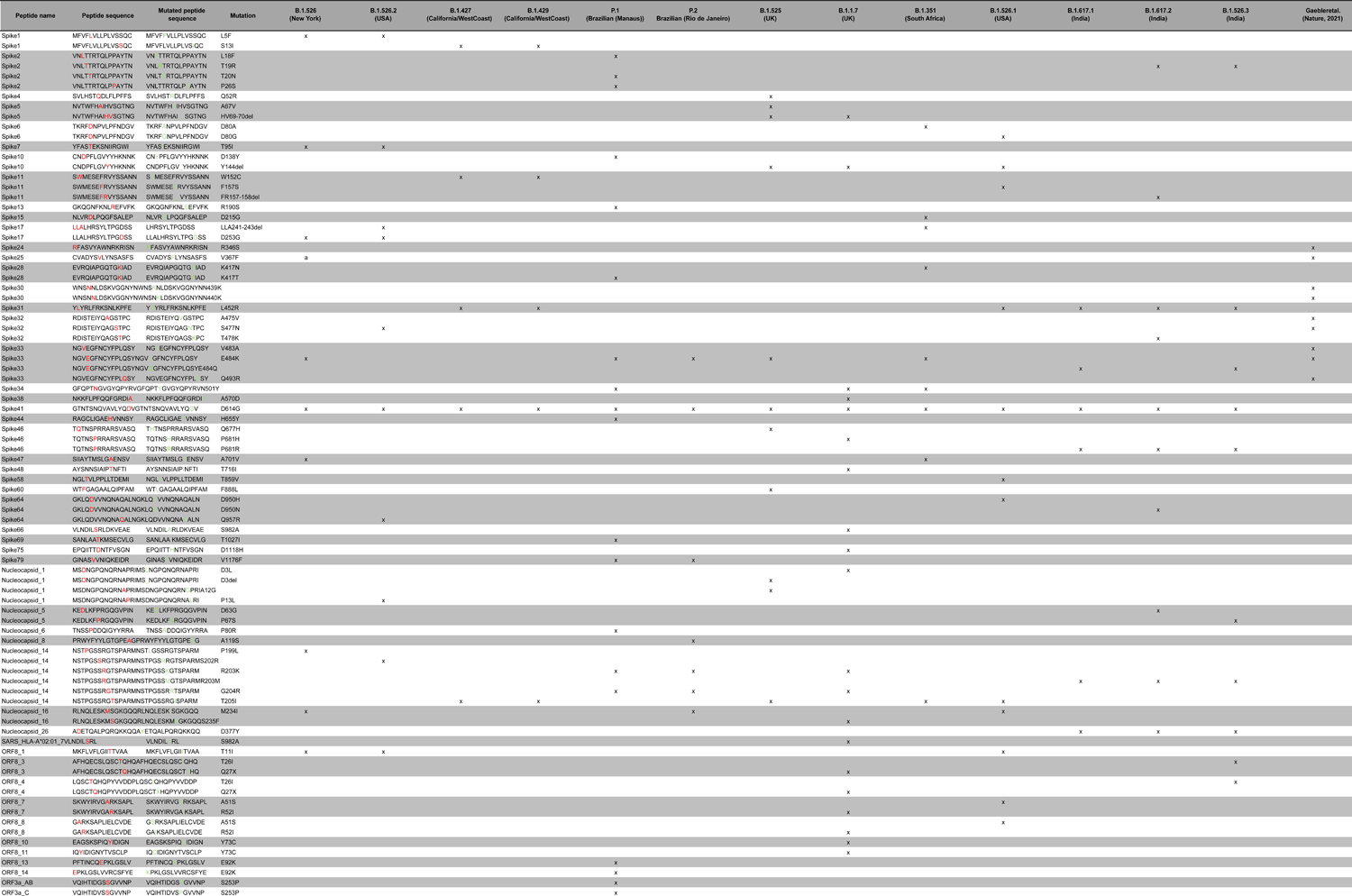
**Mutational hotspots in the peptide residues.**

**Table S10.**
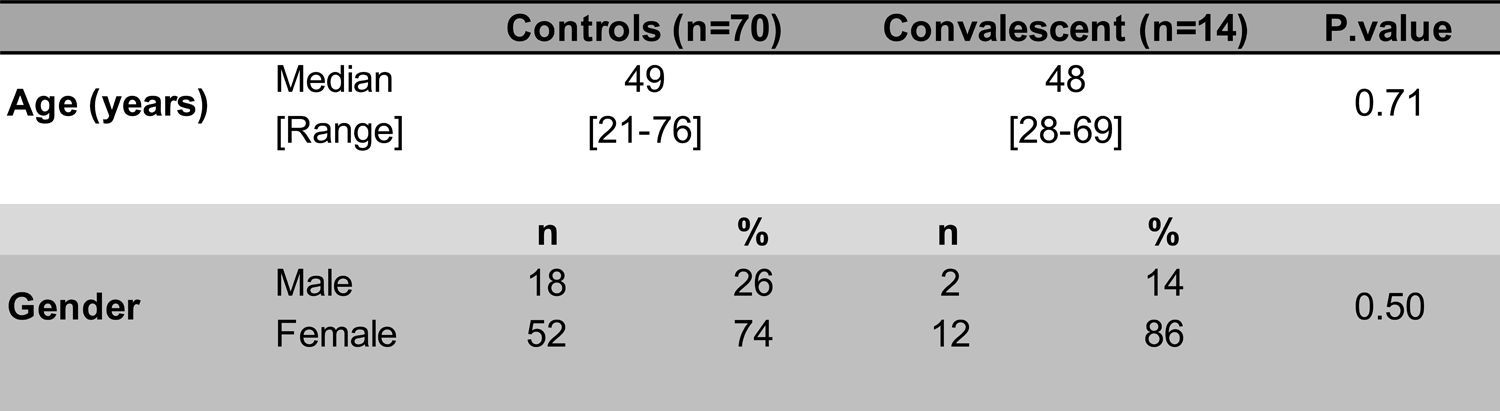
Characteristics of vaccinated healthcare workers.

## References

1. Chen, Z. & John Wherry, E. T cell responses in patients with COVID-19. Nat Rev Immunol 20, 529–536 (2020).

2. Baden, L. R. et al. Efficacy and Safety of the mRNA-1273 SARS-CoV-2 Vaccine. N Engl J Med 384, 403–416 (2021).

3. Polack, F. P. et al. Safety and Efficacy of the BNT162b2 mRNA Covid-19 Vaccine. N Engl J Med 383, 2603–2615 (2020).

4. Walsh, E. E. et al. Safety and Immunogenicity of Two RNA-Based Covid-19 Vaccine Candidates. N Engl J Med 383, 2439–2450 (2020).

5. Bilich, T. et al. T cell and antibody kinetics delineate SARS-CoV-2 peptides mediating long-term immune responses in COVID-19 convalescent individuals. Sci. Transl. Med. 13, eabf7517 (2021).

6. Garcia-Beltran, W. F. et al. Multiple SARS-CoV-2 variants escape neutralization by vaccine-induced humoral immunity. http://medrxiv.org/lookup/doi/10.1101/2021.02.14.21251704 (2021) doi:10.1101/2021.02.14.21251704.

7. Moore, J. P. Approaches for Optimal Use of Different COVID-19 Vaccines: Issues of Viral Variants and Vaccine Efficacy. JAMA 325, 1251 (2021).

8. Channappanavar, R., Fett, C., Zhao, J., Meyerholz, D. K. & Perlman, S. Virus-Specific Memory CD8 T Cells Provide Substantial Protection from Lethal Severe Acute Respiratory Syndrome Coronavirus Infection. Journal of Virology 88, 11034–11044 (2014).

9. Le Bert, N. et al. SARS-CoV-2-specific T cell immunity in cases of COVID-19 and SARS,and uninfected controls. Nature 584, 457–462 (2020). 1.

10. Ng, O.-W. et al. Memory T cell responses targeting the SARS coronavirus persist up to 11 years post-infection. Vaccine 34, 2008–2014 (2016).

11. Zhao, J. et al. Airway Memory CD4 + T Cells Mediate Protective Immunity against Emerging Respiratory Coronaviruses. Immunity 44, 1379–1391 (2016).

12. Mosmann, T. R. & Coffman, R. L. TH1 and TH2 Cells: Different Patterns of Lymphokine Secretion Lead to Different Functional Properties. Annu. Rev. Immunol. 7, 145–173 (1989).

13. Romagnani, S. Biology of human TH1 and TH2 cells. J Clin Immunol 15, 121–129 (1995).

14. Romagnani, S. & Maggi, E. Th1 versus Th2 responses in AIDS. Current Opinion in Immunology 6, 616–622 (1994).

15. Ruterbusch, M., Pruner, K. B., Shehata, L. & Pepper, M. In Vivo CD4 ^+^ T Cell Differentiation and Function: Revisiting the Th1/Th2 Paradigm. Annu. Rev. Immunol. 38, 705–725 (2020).

16. Dan, J. M. et al. Immunological memory to SARS-CoV-2 assessed for up to 8 months after infection. Science 371, eabf4063 (2021).

17. Habel, J. R. et al. Suboptimal SARS-CoV-2−specific CD8 ^+^ T cell response associated with the prominent HLA-A*02:01 phenotype. Proc Natl Acad Sci USA 117, 24384–24391 (2020).

18. Oxford Immunology Network Covid-19 Response T cell Consortium et al. Broad and strong memory CD4+ and CD8+ T cells induced by SARS-CoV-2 in UK convalescent individuals following COVID-19. Nat Immunol 21, 1336–1345 (2020).

19. Rodda, L. B. et al. Functional SARS-CoV-2-Specific Immune Memory Persists after Mild COVID-19. Cell 184, 169–183.e17 (2021).

20. Sekine, T. et al. Robust T Cell Immunity in Convalescent Individuals with Asymptomatic or Mild COVID-19. Cell 183, 158–168.e14 (2020).

21. Bacher, P. et al. Low-Avidity CD4+ T Cell Responses to SARS-CoV-2 in Unexposed Individuals and Humans with Severe COVID-19. Immunity 53, 1258–1271.e5 (2020).

22. Lee, J.-S. et al. Evidence of Severe Acute Respiratory Syndrome Coronavirus 2 Reinfection After Recovery from Mild Coronavirus Disease 2019. Clinical Infectious Diseases ciaa1421 (2020) doi:10.1093/cid/ciaa1421.

23. Braun, J. et al. SARS-CoV-2-reactive T cells in healthy donors and patients with COVID-19. Nature 587, 270–274 (2020).

24. Grifoni, A. et al. Targets of T Cell Responses to SARS-CoV-2 Coronavirus in Humans with COVID-19 Disease and Unexposed Individuals. Cell 181, 1489–1501.e15 (2020).

25. Dykema, A. G. et al. Functional characterization of CD4+ T cell receptors crossreactive for SARS-CoV-2 and endemic coronaviruses. Journal of Clinical Investigation 131, e146922 (2021).

26. Goubet, A.-G. et al. Prolonged SARS-CoV-2 RNA virus shedding and lymphopenia are hallmarks of COVID-19 in cancer patients with poor prognosis. http://medrxiv.org/lookup/doi/10.1101/2021.04.26.21250357 (2021) doi:10.1101/2021.04.26.21250357.

27. Andre, F. et al. Malignant effusions and immunogenic tumour-derived exosomes. The Lancet 360, 295–305 (2002).

28. Besse, B. et al. Dendritic cell-derived exosomes as maintenance immunotherapy after first line chemotherapy in NSCLC. OncoImmunology 5, e1071008 (2016).

29. Borg, C. et al. Novel mode of action of c-kit tyrosine kinase inhibitors leading to NK cell–dependent antitumor effects. J. Clin. Invest. 114, 379–388 (2004).

30. Wemeau, M. et al. Calreticulin exposure on malignant blasts predicts a cellular anticancer immune response in patients with acute myeloid leukemia. Cell Death Dis 1, e104–e104 (2010).

31. Mateus, J. et al. Selective and cross-reactive SARS-CoV-2 T cell epitopes in unexposed humans. Science 370, 89–94 (2020).

32. Nelde, A. et al. SARS-CoV-2-derived peptides define heterologous and COVID-19-induced T cell recognition. Nat Immunol 22, 74–85 (2021).

33. Le Bert, N. et al. Highly functional virus-specific cellular immune response in asymptomatic SARS-CoV-2 infection. Journal of Experimental Medicine 218, e20202617 (2021).

34. Zhao, Y. et al. Clonal expansion and activation of tissue-resident memory-like T H 17 cells expressing GM-CSF in the lungs of patients with severe COVID-19. Sci. Immunol. 6, eabf6692 (2021).

35. Fournier, P.-E. et al. Emergence and outcomes of the SARS-CoV-2 ‘Marseille-4’ variant. International Journal of Infectious Diseases 106, 228–236 (2021).

36. Cha, E. et al. Improved Survival with T Cell Clonotype Stability After Anti-CTLA-4 Treatment in Cancer Patients. Science Translational Medicine 6, 238ra70-238ra70 (2014).

37. Scheper, W. et al. Low and variable tumor reactivity of the intratumoral TCR repertoire in human cancers. Nat Med 25, 89–94 (2019).

38. Yager, E. J. et al. Age-associated decline in T cell repertoire diversity leads to holes in the repertoire and impaired immunity to influenza virus. Journal of Experimental Medicine 205, 711–723 (2008).

39. Shang, J. et al. Structural basis of receptor recognition by SARS-CoV-2. Nature 581, 221–224 (2020).

40. Low, J. S. et al. Clonal analysis of immunodominance and cross-reactivity of the CD4 T cell response to SARS-CoV-2. Science eabg8985 (2021)

41. Gaebler, C. et al. Evolution of antibody immunity to SARS-CoV-2. Nature 591, 639– 644 (2021).

42. Mouton, W. et al. A novel whole-blood stimulation assay to detect and quantify memory T-cells in COVID-19 patients. http://medrxiv.org/lookup/doi/10.1101/2021.03.11.21253202 (2021) doi:10.1101/2021.03.11.21253202.

43. Whitmire, J. K., Asano, M. S., Murali-Krishna, K., Suresh, M. & Ahmed, R. Long-Term CD4 Th1 and Th2 Memory following Acute Lymphocytic Choriomeningitis Virus Infection. J Virol 72, 8281–8288 (1998).

44. Hondowicz, B. D., Kim, K. S., Ruterbusch, M. J., Keitany, G. J. & Pepper, M. IL-2 is required for the generation of viral-specific CD4 ^+^ Th1 tissue-resident memory cells and B cells are essential for maintenance in the lung. Eur. J. Immunol. 48, 80–86 (2018).

45. Li, C. K. et al. T Cell Responses to Whole SARS Coronavirus in Humans. J Immunol 181, 5490–5500 (2008).

46. Page, C. et al. Induction of Alternatively Activated Macrophages Enhances Pathogenesis during Severe Acute Respiratory Syndrome Coronavirus Infection. Journal of Virology 86, 13334–13349 (2012).

47. Yale IMPACT Team et al. Longitudinal analyses reveal immunological misfiring in severe COVID-19. Nature 584, 463–469 (2020).

48. Kumamoto, Y. et al. CD301b+ Dermal Dendritic Cells Drive T Helper 2 Cell-Mediated Immunity. Immunity 39, 733–743 (2013).

49. Sokol, C. L., Camire, R. B., Jones, M. C. & Luster, A. D. The Chemokine Receptor CCR8 Promotes the Migration of Dendritic Cells into the Lymph Node Parenchyma to Initiate the Allergic Immune Response. Immunity 49, 449–463.e6 (2018).

50. Van Dyken, S. J. et al. A tissue checkpoint regulates type 2 immunity. Nat Immunol 17, 1381–1387 (2016).

51. Sánchez-Cerrillo, I. et al. COVID-19 severity associates with pulmonary redistribution of CD1c+ DCs and inflammatory transitional and nonclassical monocytes. Journal of Clinical Investigation 130, 6290–6300 (2020).

52. Becattini, S. et al. Functional heterogeneity of human memory CD4+ T cell clones primed by pathogens or vaccines. Science 347, 400–406 (2015).

53. Honda, T. et al. Tuning of Antigen Sensitivity by T Cell Receptor-Dependent Negative Feedback Controls T Cell Effector Function in Inflamed Tissues. Immunity 40, 235– 247 (2014).

54. Shinoda, K. et al. Thy1 ^+^ IL-7 ^+^ lymphatic endothelial cells in iBALT provide a survival niche for memory T-helper cells in allergic airway inflammation. Proceedings of the National Academy of Sciences 113, E2842–E2851 (2016).

55. Yeon, S. et al. IL-7 plays a critical role for the homeostasis of allergen-specific memory CD4 T cells in the lung and airways. Scientific Reports 7, (2017).

56. Meckiff, B. J. et al. Imbalance of Regulatory and Cytotoxic SARS-CoV-2-Reactive CD4+ T Cells in COVID-19. Cell 183, 1340–1353.e16 (2020).

57. Weiskopf, D. et al. Phenotype and kinetics of SARS-CoV-2-specific T cells in COVID-19 patients with acute respiratory distress syndrome. Science Immunology 5, eabd2071 (2020).

58. Bacher, P. et al. Human Anti-fungal Th17 Immunity and Pathology Rely on Cross-Reactivity against Candida albicans. Cell 176, 1340–1355.e15 (2019).

59. Greiling, T. M. et al. Commensal orthologs of the human autoantigen Ro60 as triggers of autoimmunity in lupus. Science Translational Medicine 10, eaan2306 (2018).

60. Koutsakos, M. et al. Human CD8+ T cell cross-reactivity across influenza A, B and C viruses. Nature Immunology 20, 613–625 (2019).

61. Sridhar, S. et al. Cellular immune correlates of protection against symptomatic pandemic influenza. Nature Medicine 19, 1305–1312 (2013).

62. Welsh, R. M., Che, J. W., Brehm, M. A. & Selin, L. K. Heterologous immunity between viruses: Heterologous immunity between viruses. Immunological Reviews 235, 244– 266 (2010).

63. Woodland, D. L. & Blackman, M. A. Immunity and age: living in the past? Trends in Immunology 27, 303–307 (2006).

64. Tarke, A. et al. Comprehensive analysis of T cell immunodominance and immunoprevalence of SARS-CoV-2 epitopes in COVID-19 cases. Cell Reports Medicine 2, 100204 (2021).

65. Tan, A. T. et al. Early induction of functional SARS-CoV-2-specific T cells associates with rapid viral clearance and mild disease in COVID-19 patients. Cell Reports 34, 108728 (2021).

66. Hansen, T. H. & Bouvier, M. MHC class I antigen presentation: learning from viral evasion strategies. Nature Reviews Immunology 9, 503–513 (2009).

67. Hachim, A. et al. ORF8 and ORF3b antibodies are accurate serological markers of early and late SARS-CoV-2 infection. Nature Immunology 21, 1293–1301 (2020).

68. Young, B. E. et al. Effects of a major deletion in the SARS-CoV-2 genome on the severity of infection and the inflammatory response: an observational cohort study. The Lancet 396, 603–611 (2020).

69. Su, Y. C. F. et al. Discovery and Genomic Characterization of a 382-Nucleotide Deletion in ORF7b and ORF8 during the Early Evolution of SARS-CoV-2. mBio 11, (2020).

70. Greaney, A. J. et al. Complete Mapping of Mutations to the SARS-CoV-2 Spike Receptor-Binding Domain that Escape Antibody Recognition. Cell Host & Microbe 29, 44–57.e9 (2021).

71. Koenig, P.-A. et al. Structure-guided multivalent nanobodies block SARS-CoV-2 infection and suppress mutational escape. Science 371, eabe6230 (2021).

72. Agerer, B. et al. SARS-CoV-2 mutations in MHC-I-restricted epitopes evade CD8 + T cell responses. Science Immunology 6, eabg6461 (2021).

73. Iwasaki, A. & Omer, S. B. Why and How Vaccines Work. Cell 183, 290–295 (2020).

74. Bolles, M. et al. A Double-Inactivated Severe Acute Respiratory Syndrome Coronavirus Vaccine Provides Incomplete Protection in Mice and Induces Increased Eosinophilic Proinflammatory Pulmonary Response upon Challenge. Journal of Virology 85, 12201–12215 (2011).

75. Tseng, C.-T. et al. Immunization with SARS Coronavirus Vaccines Leads to Pulmonary Immunopathology on Challenge with the SARS Virus. PLoS ONE 7, e35421 (2012).

76. Jiang, S. et al. Roadmap to developing a recombinant coronavirus S protein receptor-binding domain vaccine for severe acute respiratory syndrome. Expert Review of Vaccines 11, 1405–1413 (2012).

77. Martins, K. A., Bavari, S. & Salazar, A. M. Vaccine adjuvant uses of poly-IC and derivatives. Expert Review of Vaccines 14, 447–459 (2015).

78. Jo, W. K., Drosten, C. & Drexler, J. F. The evolutionary dynamics of endemic human coronaviruses. Virus Evolution 7, (2021).

79. Chaput, N. et al. Phase I clinical trial combining imatinib mesylate and IL-2: HLA-DR ^+^ NK cell levels correlate with disease outcome. OncoImmunology 2, e23080 (2013).

80. Fournier, P.-E. et al. Emergence and outcomes of the SARS-CoV-2 ‘Marseille-4’ variant. International Journal of Infectious Diseases 106, 228–236 (2021).

81. Sahin, U. et al. BNT162b2 vaccine induces neutralizing antibodies and poly-specific T cells in humans. Nature (2021) doi:10.1038/s41586-021-03653-6.

82. Saini, S. K. et al. SARS-CoV-2 genome-wide T cell epitope mapping reveals immunodominance and substantial CD8 + T cell activation in COVID-19 patients. Science Immunology 6, eabf7550 (2021).

83. Shomuradova, A. S. et al. SARS-CoV-2 Epitopes Are Recognized by a Public and Diverse Repertoire of Human T Cell Receptors. Immunity 53, 1245–1257.e5 (2020).

